# Estimating local outbreak risks and the effects of non-pharmaceutical interventions in age-structured populations: SARS-CoV-2 as a case study

**DOI:** 10.1101/2021.04.27.21256163

**Authors:** Francesca A. Lovell-Read, Silvia Shen, Robin N. Thompson

**Author notes:** Corresponding author Address: Merton College, Merton Street, Oxford, OX1 4JD.

## Abstract

During the COVID-19 pandemic, non-pharmaceutical interventions (NPIs) including school closures, workplace closures and social distancing policies have been employed worldwide to reduce transmission and prevent local outbreaks. However, transmission and the effectiveness of NPIs depend strongly on age-related factors including heterogeneities in contact patterns and pathophysiology. Here, using SARS-CoV-2 as a case study, we develop a branching process model for assessing the risk that an infectious case arriving in a new location will initiate a local outbreak, accounting for the age distribution of the host population. We show that the risk of a local outbreak depends on the age of the index case, and we explore the effects of NPIs targeting individuals of different ages. Social distancing policies that reduce contacts outside of schools and workplaces and target individuals of all ages are predicted to reduce local outbreak risks substantially, whereas school closures have a more limited impact. In the scenarios considered here, when different NPIs are used in combination the risk of local outbreaks can be eliminated. We also show that heightened surveillance of infectious individuals reduces the level of NPIs required to prevent local outbreaks, particularly if enhanced surveillance of symptomatic cases is combined with efforts to find and isolate nonsymptomatic infected individuals. Our results reflect real-world experience of the COVID-19 pandemic, during which combinations of intense NPIs have reduced transmission and the risk of local outbreaks. The general modelling framework that we present can be used to estimate local outbreak risks during future epidemics of a range of pathogens, accounting fully for age-related factors.

## 1. Introduction

Throughout the COVID-19 pandemic, policy makers worldwide have relied on non-pharmaceutical interventions (NPIs) to limit the spread of SARS-CoV-2. Commonly introduced NPIs have included school closures, workplace closures and population-wide social distancing policies, all of which aim to reduce the numbers of contacts between individuals and disrupt potential chains of transmission [1–4]. Similar measures have previously been adopted for countering other infectious diseases such as Ebola and pandemic influenza [5–7], and are likely to remain a key line of defence against emerging pathogens that are directly transmitted between hosts. NPIs are particularly important when no effective treatment or vaccine is available, and they are also beneficial when vaccination programmes are being rolled out [8–10]. If vaccines do not prevent transmission completely, then NPIs may be important even when vaccination is widespread [11]. However, the negative economic, social and non-disease health consequences of NPIs have been widely discussed, with the impact of school closures on the academic progress and wellbeing of school-aged individuals a particular concern [7, 12–16]. Therefore, assessing the effectiveness of different NPIs at reducing transmission is critical for determining whether or not they should be used.

Since NPIs such as school and workplace closures affect distinct age groups within the population, when evaluating their effectiveness it is important to account for age-dependent factors that influence transmission. Multiple studies have documented marked heterogeneities in the patterns of contacts between individuals in different age groups, with school-aged individuals tending to have more contacts each day than older individuals [17–23]. Since close contact between individuals is a key driver of transmission for respiratory pathogens such as influenza viruses and SARS-CoV-2, these contact patterns influence transmission dynamics and consequently the effects of interventions that target different age groups [18, 19, 24–27]. Additionally, many diseases are characterised by significant age-related variations in pathophysiology. For example, for SARS-CoV-2, children may be less susceptible to infection than adults [27–31], and more likely to experience asymptomatic or subclinical courses of infection [28, 31–36]. Since the secondary attack rate (the proportion of close contacts that lead to new infections) from asymptomatic or subclinical hosts is lower than from hosts with clinical symptoms [37–42], children are likely to be less infectious on average than older individuals who are at increased risk of developing symptoms [43–45].

Previous studies have used age-stratified deterministic transmission models to investigate the effects of NPIs on COVID-19 epidemic peak incidence and timing. Prem *et al.* [46] projected the outbreak in Wuhan, China, over a one year period under different control scenarios, and demonstrated that a period of intense control measures including school closures, a 90% reduction in the workforce and a significant reduction in other social mixing could delay the epidemic peak by several months. Zhang *et al.* [27] predicted that eliminating all school contacts during the outbreak period would lead to a noticeable decrease in the peak incidence and a later peak; however, they did not take differences between symptomatic and asymptomatic cases into account explicitly. In contrast, Davies *et al.* [31] used estimates of age-dependent susceptibility and clinical fraction fitted to the observed age distribution of cases in six countries to demonstrate that school closures alone were unlikely to reduce SARS-CoV-2 transmission substantially. Davies *et al.* [47] subsequently concluded that a combination of several strongly enforced NPIs would be necessary to avoid COVID-19 cases exceeding available healthcare capacity in the UK.

Rather than considering the entire epidemic curve, here we focus on estimating the probability that cases introduced to a new location trigger a local outbreak as opposed to fading out with few cases. Localised clusters of transmission have been a feature of the COVID-19 pandemic [48–50], and assessing the risk that such local outbreaks occur requires a stochastic model in which the pathogen can either invade or fade out. Stochastic branching process models have been applied previously to assess outbreak risks for many pathogens without considering different age groups explicitly [51–55], and extended to consider adults and children as two distinct groups [56]. However, the significant heterogeneities in contact patterns and pathophysiology between individuals across the full range of ages have never previously been considered in estimates of local outbreak risks. Here, we develop an age-structured branching process model that can be used to estimate the probability of a local outbreak occurring for index cases of different ages, and demonstrate how the age-dependent risk profile changes when susceptibility to infection and clinical fraction vary with age.

We use the model to investigate the effects on the local outbreak probability of NPIs that reduce the numbers of contacts between individuals. Specifically, we use location-specific contact data for the UK detailing the average numbers of daily contacts occurring in school, in the workplace and elsewhere [17] to model the impacts of school closures, workplace closures and broader social distancing policies. We demonstrate that, for SARS-CoV-2, contacts occurring outside schools and the workplace are a key driver of sustained transmission. Thus, population-wide social distancing policies that affect individuals of all ages lead to a substantial reduction in the risk of local outbreaks. In contrast, since school-aged individuals only make up around one quarter of the UK population and tend to have large numbers of contacts outside school, school closures are predicted to have only a limited effect when applied as the sole NPI.

We then go on to consider the impacts of mixed strategies made up of multiple NPIs, as well as additional NPIs that do not only reduce numbers of contacts. Specifically, we show that rigorous surveillance and effective isolation of infected hosts can reduce the level of contact-reducing NPIs required to achieve substantial reductions in the risk of local outbreaks. Although we use SARS-CoV-2 as a case study, our approach can be applied more generally to explore the effects of NPIs on the risk of outbreaks of any pathogen for which age-related heterogeneities play a significant role in transmission dynamics.

## 2. Methods

### 2.1 Mathematical model

We considered a branching process model in which the population was divided into 16 age groups, denoted *G*_1_, *G*_2_, … , *G*_1#_. The first 15 groups represent individuals aged 0-74, divided into five-year intervals (0-4, 5-9, 10-14 etc.). The final group represents individuals aged 75 and over. The total number of individuals in age group *G*_*k*_ is denoted *N*_*k*_. Infected individuals in each age group *G*_*k*_ are classified into compartments representing asymptomatic (*A*_*k*_), presymptomatic (*P*_*k*_) or symptomatic (*S*_*k*_) hosts, where an individual in the *A*_*k*_ compartment does not develop symptoms at any time during their course of infection.

An infected individual of any type in group *G*_*k*_ may generate new infections in any age group. In our model, the rate at which a single infected symptomatic individual in group *G*_*k*_ generates infections in group *G*_j_ is given by

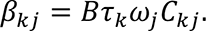

Here, τ_*k*_ represents the infectivity of individuals in group *G*_*k*_, ω_j_ represents the susceptibility to infection of individuals in group *G*_j_, *C*_*k*j_ represents the daily number of unique contacts a single individual in group *G*_*k*_ has with individuals in group *G*_j_, and *B* is a scaling factor that can be used to set the reproduction number of the pathogen being considered (see Section 2.2). Since the initial phase of potential local outbreaks are the focus of this study, we did not account for depletion of susceptible hosts explicitly. The relative transmission rates from presymptomatic and asymptomatic individuals compared to symptomatic individuals are given by the scaled quantities ηβ_*k*j_and *θβ*_*k*j_, respectively, where *η* and *θ* were chosen so that the proportions of transmissions generated by presymptomatic and asymptomatic hosts were in line with literature estimates [57]. The parameter *ξ*_*k*_ represents the proportion of asymptomatic infections in group *G*_*k*_, so that a new infection in group *G*_*k*_ either increases *A*_*k*_ by one (with probability *ξ*_*k*_) or increases *P*_*k*_ by one (with probability 1 − *ξ*_*k*_).

A presymptomatic individual in group *G*_*k*_ may go on to develop symptoms (transition from *P*_*k*_ to *S*_*k*_) or be detected and isolated (so that *P*_*k*_ decreases by one). A symptomatic individual in group *G*_*k*_ may be detected and isolated as a result of successful surveillance, or may be removed due to self-isolation, recovery or death (so that *S*_*k*_ decreases by one in either case). Similarly, an asymptomatic individual in group *G*_*k*_ may be detected and isolated or recover (so that *A*_*k*_ decreases by one). A schematic of the different possible events in the model is shown in Fig 1.

**Fig 1.**
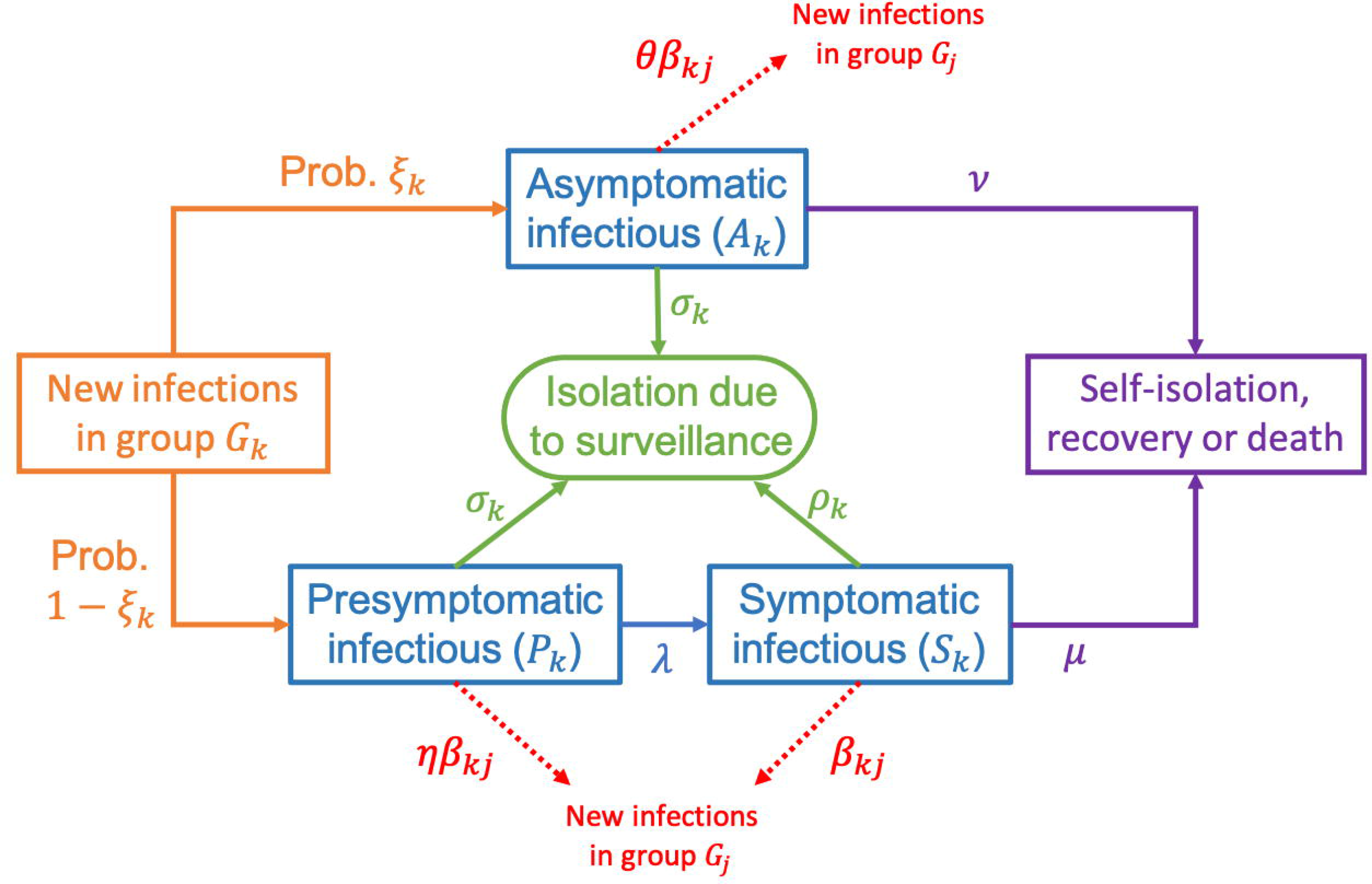
The branching process model used in our analyses. Schematic showing the different possible events in the branching process model and the rates at which they occur. The parameters of the model are described in the text and in Tables 1, 2 and 3.

**Table 1.**
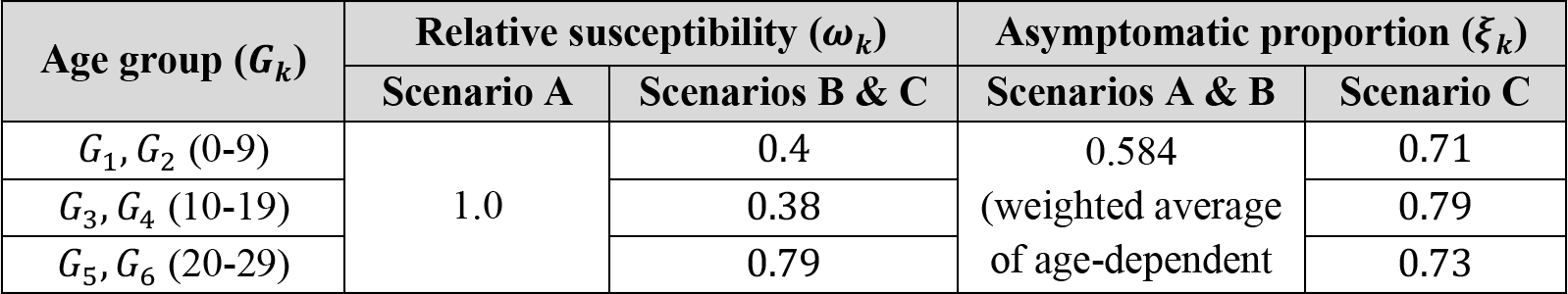
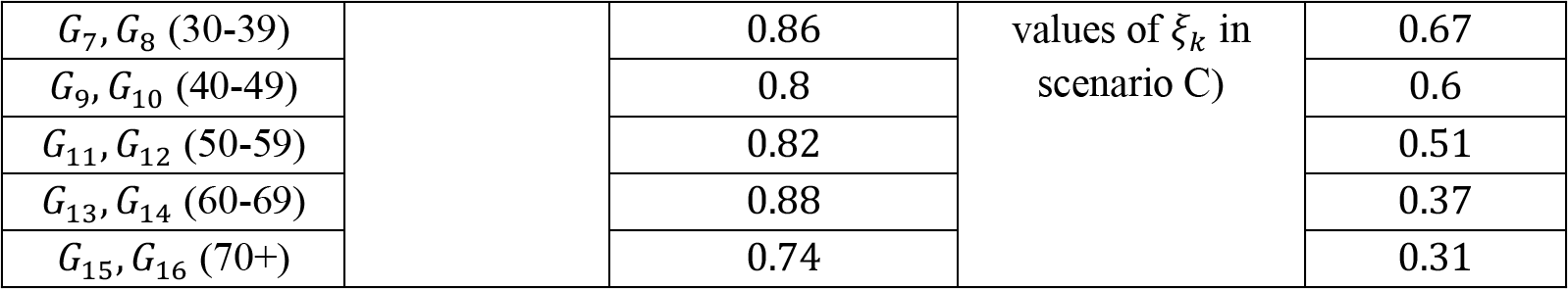
Baseline values of age-dependent parameters. Values used for the age-dependent relative susceptibility to infection (ω_*k*_) and the proportion of infections that are asymptomatic (*ξ*_*k*_) for each of the scenarios A, B and C [31].

**Table 2.** Baseline values of scenario-independent parameters. Values used for the parameters that were assumed not to vary between scenarios A, B and C. We also considered strategies involving enhanced surveillance (*ρ*_*k*_, *σ*_*k*_ > 0) – see Fig 6.

| Parameter | Meaning | Baseline value | Justification |
| --- | --- | --- | --- |
| $R_0$ | Expected number of secondary infections generated by a single infectious host in the absence of interventions | $R_0 = 3$ | Within estimated range for original SARS-CoV-2 virus [63-66] |
| $\lambda$ | Rate at which presymptomatic hosts develop symptoms | $\lambda = 1/2 \text{ days}^{-1}$ | [59] |
| $\mu$ | Rate at which symptomatic hosts are removed due to self-isolation, recovery or death | $\mu = 1/8 \text{ days}^{-1}$ | [60-62] |
| $\nu$ | Rate at which asymptomatic hosts are removed due to recovery or death | $\nu = 1/10 \text{ days}^{-1}$ | Chosen so that, in the absence of interventions, the expected duration of infection is identical for all infected hosts ($\frac{1}{\nu} = \frac{1}{\lambda} + \frac{1}{\mu}$) |
| $\tau_k$ | (Relative) infectivity parameter | $\tau_k = 1$ for $k = 1, \dots, 16$ | Assumed |
| $\rho_k$ | Isolation rate due to surveillance of symptomatic individuals | $\rho_k = 0$ for $k = 1, \dots, 16$ | N/A |
| $\sigma_k$ | Isolation rate due to surveillance of nonsymptomatic individuals | $\sigma_k = 0$ for $k = 1, \dots, 16$ | N/A |

**Table 3.** Baseline values of scenario-dependent scaling parameters. Values used for the scaling parameters *B*, *η* and *θ* for each of the scenarios A, B and C.

| Parameter | Meaning | Value |  |  | Justification |
| --- | --- | --- | --- | --- | --- |
|  |  | Scenario A | Scenario B | Scenario C |  |
| $B$ | Transmission rate scaling factor | 0.0386 | 0.0559 | 0.0607 | Chosen so that $R_0 = 3$ [63-66] |
| $\eta$ | Relative transmission rate of presymptomatic hosts compared to symptomatic hosts | 4.83 | 4.83 | 4.83 | Chosen so that the proportion of all infections arising from presymptomatic hosts is 0.489 [57] |
| $\theta$ | Relative transmission rate of asymptomatic hosts compared to symptomatic hosts | 0.149 | 0.149 | 0.130 | Chosen so that the proportion of all infections arising from entirely asymptomatic hosts is 0.106 [57] |

The parameter *λ* represents the rate at which presymptomatic individuals develop symptoms, so that the expected duration of the presymptomatic infectious period is 1/*λ* days in the absence of surveillance of nonsymptomatic infected individuals. Similarly, the expected duration of the asymptomatic infectious period in the absence of surveillance is 1/*ν* days. The parameter *μ* represents the rate at which symptomatic individuals are removed as a result of self-isolation, recovery or death, so that the duration of time for which they are able to infect others is 1/*μ* days.

For each group *G*_*k*_, the rate at which symptomatic individuals are detected and isolated as a result of enhanced surveillance is determined by the parameter *ρ*_*k*_. Analogously, the parameter *σ*_*k*_ governs the rate at which presymptomatic and asymptomatic individuals in *G*_*k*_ are detected and isolated. We assumed that surveillance measures targeting nonsymptomatic hosts are equally effective for those who are presymptomatic and those who are asymptomatic, and therefore used the same rate of isolation due to surveillance for both of these groups.

### 2.2 Reproduction number

The effective reproduction number, *R*, represents the expected number of secondary infections generated by a single infected individual during their entire course of infection, accounting for interventions that are in place. Here, we take a heuristic approach to derive the following expression for *R*:

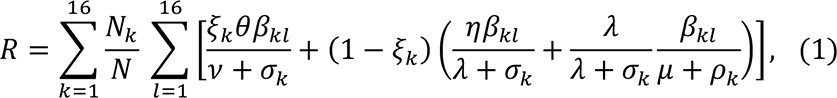

where *N* = *N*_1_ + ⋯ + *N*_1#_ is the total population size. To obtain this expression, we first consider the expected number of secondary infections an infected individual in age group *G*_*k*_ will generate in age group *G*_*l*_. If an individual in age group *G*_*k*_ experiences a fully asymptomatic course of infection, which occurs with probability *ξ*_*k*_, they will generate new infections in age group *G*_*l*_ at rate *θβ*_*kl*_ and recover or be isolated at rate *λ* + *σ*_*k*_. Therefore, the total number of infections they are expected to cause in age group *G*_*l*_ is *θβ*_*kl*_/(*λ* + *σ*_*k*_). If instead the individual in age group *G*_*k*_ experiences a symptomatic course of infection, which occurs with probability 1 − *ξ*_*k*_, whilst presymptomatic they will generate new infections in age group *G*_*l*_ at rate *ηβ*_*kl*_ and be isolated or develop symptoms at rate *λ* + *σ*_*k*_. Thus, the total number of infections they are expected to cause in age group *G*_*l*_ whilst presymptomatic is *ηβ*_*kl*_/(*λ* + *σ*_*k*_). If they go on to develop symptoms before being isolated, which occurs with probability *λ*/(*λ* + *σ*_*k*_), applying similar reasoning they are expected to cause β_*kl*_/(*μ* + *ρ*_*k*_) new infections in age group *G*_*l*_ whilst symptomatic.

Combining these possibilities leads to the term in square brackets in expression (1), which is then summed over all possible age groups *G*_*l*_ of the infectee. Finally, to obtain the full expression (1) we take a weighted average across all possible age groups *G*_*k*_ of the infector, where the weights *N*_*k*_/*N* represent the proportions of the population belonging to each age group. This corresponds to the assumption that the initial infected host is more likely to belong to an age group containing more individuals than an age group with fewer individuals.

In the absence of interventions, i.e. when *σ*_*k*_ = *ρ*_*k*_ = 0 (representing no enhanced isolation as a result of surveillance) and β_*kl*_ is calculated using contact patterns that are characteristic of normal behaviour, the effective reproduction number, *R*, is equal to the basic reproduction number, *R*_(_.

### 2.3 Model parameterisation

The numbers of individuals in each age group (values of *N*_*k*_) were chosen according to United Nations age demographic data for the UK [58] (Fig 2A). The daily numbers of contacts between individuals in each age group (values of *C*_*k*j_) were set according to the 16x16 ‘contact matrix’ for the UK, in which the (*k*, *j*)th entry represents the expected daily number of unique contacts an individual in age group *G*_*k*_ has with individuals in age group *G*_j_ [17]. In addition to matrices representing ‘all’ contacts (Figure 2B), we also considered matrices detailing a breakdown into ‘school’, ‘work’, ‘home’ and ‘other’ contacts (Figures 2C-F), allowing us to investigate the effects of control interventions that reduce contacts in each of these settings.

**Fig 2.**
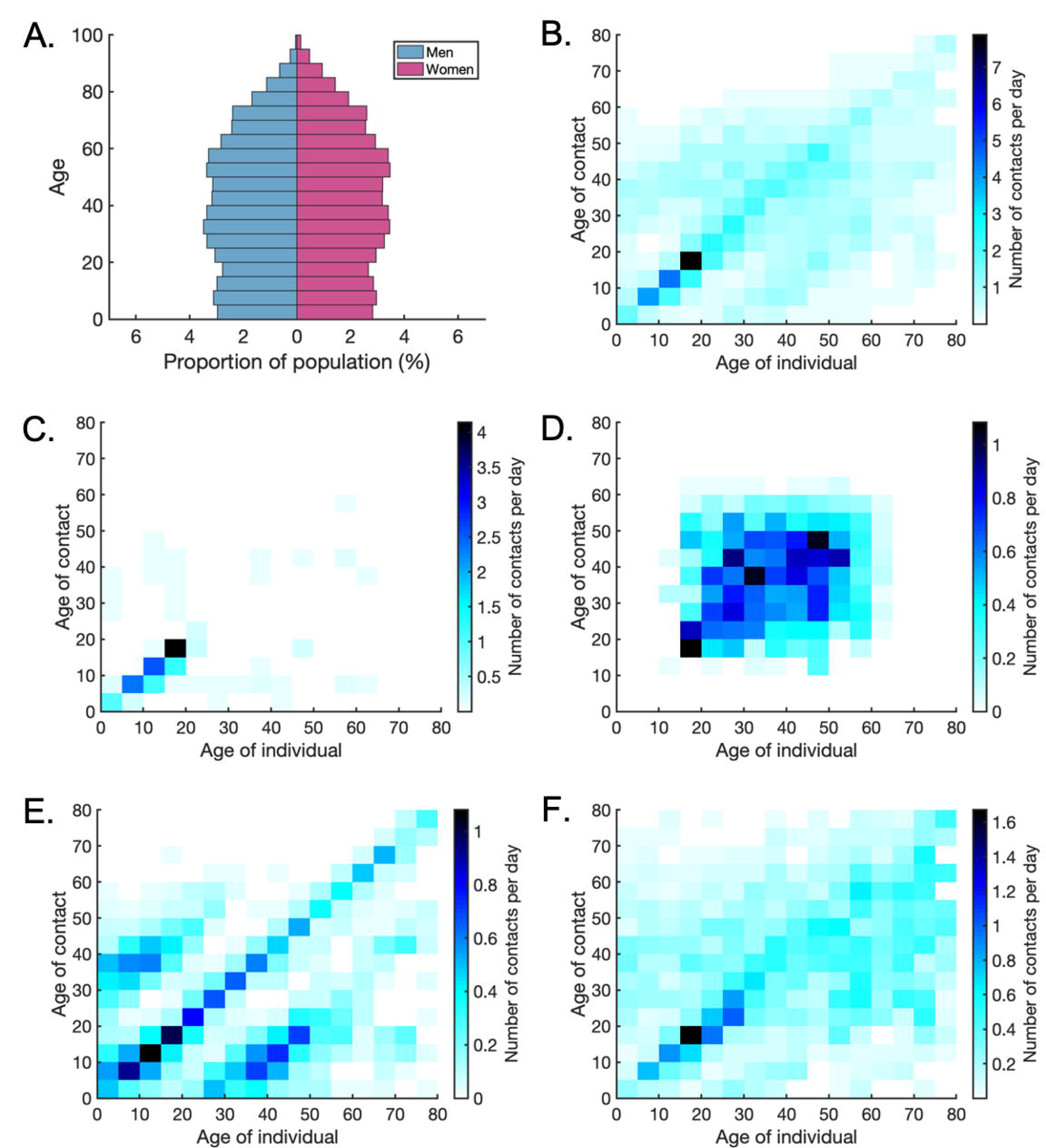
Age demographic and age-structured contact patterns for the United Kingdom. A. United Nations age demographic data for the UK in 2020, split into five-year age groups (where the final age group contains all ages 75+) [58]. B. Heat map of UK ‘all contacts’ matrix, representing the expected daily number of unique contacts that an individual in each age group *G*_*k*_ has with individuals in each other age group *G*_&_ [17]. C. The analogous figure to B, but showing only the subset of ‘all’ contacts that occur in schools (‘school’ contacts). D. The analogous figure to C, but showing only ‘workplace’ contacts. E. The analogous figure to C, but showing only ‘home’ contacts. F. The analogous figure to C, but showing only ‘other’ contacts (i.e. all contacts outside schools, workplaces or homes).

Since we considered SARS-CoV-2 as a case study, we used studies conducted during the COVID-19 pandemic to inform the epidemiological parameters of our model. Despite previous research assessing the relationships between age and factors such as susceptibility to SARS-CoV-2 infection or the propensity to develop symptoms, there is some variation in estimated parameters between different studies. To test the robustness of our results to this uncertainty, we conducted our analyses under three different scenarios (A, B and C). In scenario A, we assumed that susceptibility to infection (values of ω_*k*_) and the proportion of hosts who experience a fully asymptomatic course of infection (values of *ξ*_*k*_) are independent of age. In scenario B, susceptibility was assumed to vary with age but the proportion of asymptomatic infections is independent of age. In scenario C, we allowed both susceptibility and the asymptomatic proportion to vary with age. The values used for the parameters ω_*k*_ and *ξ*_*k*_ in each of these three scenarios are shown in Table 1 (see also [31]).

In all scenarios considered, the inherent infectivity was not assumed to be age-dependent (i.e. τ_*k*_ = 1 for all values of *k*). In other words, the expected infectiousness of infected hosts in different age groups was governed solely by the proportion of asymptomatic infections in that age group. We chose the scaling factors *η* and *θ* for the relative transmission rates from presymptomatic and asymptomatic individuals compared to symptomatic individuals so that the proportions of infections arising from each of these groups were in line with literature estimates (see Table 3 and [57]).

In the absence of enhanced isolation, we set the expected duration of the presymptomatic infectious period and the time for which symptomatic individuals are able to infect others to be 1/*λ* = 2 days and 1/*μ* = 8 days, respectively [59–62]. The asymptomatic infectious period was then chosen so that all infected individuals are expected to be infectious for the same period (i.e. 1/*λ* = 10 days). In our initial analysis, we set the isolation rates *ρ*_*k*_ and *σ*_*k*_ equal to 0 for all *k*; later, we considered the effects of increasing these rates.

Initially, we fixed *R*_(_ = 3 (in line with initial estimates of SARS-CoV-2 transmissibility [63–66], before the emergence of more transmissible variants) and used expression (1) to determine the appropriate corresponding value of the scaling factor *B*. Later, when considering the impact of NPIs on the probability of a local outbreak, we retained this value of *B* and used expression (1) to determine how the reproduction number changes as a result of the control implemented.

### 2.4 Probability of a local outbreak

The probability that an infected individual in a particular age group initiates a local outbreak when they are introduced into the population was calculated using the branching process model. One possible approach for approximating the age-dependent local outbreak probability using a branching process model is to run a large number of stochastic simulations of the model starting from a single infected individual in a particular age group, and record the proportion of simulations in which the pathogen does not fade out after only a small number of infections [67]. This would then need to be repeated for index cases of different ages. Here, we instead take an analytic approach, and derive a nonlinear system of simultaneous equations that determine the age-dependent outbreak probabilities, as described below. The local outbreak probabilities are then obtained by solving these equations numerically, and are analogous to the probabilities that would be derived from the simulation approach in the limit of infinitely many simulations. The benefit of our analytic approach is that it does not require a large number of stochastic simulations to be run.

The probability of a local outbreak not occurring (i.e. pathogen fadeout occurs), starting from a single symptomatic (or presymptomatic, asymptomatic respectively) infectious individual in age group *G*_*k*_, was denoted by *x*_*k*_ (*y*_*k*_, *z*_*k*_). Beginning with a single symptomatic individual in *G*_*k*_, the possibilities for the next event are as follows:

1. The infected individual in *G*_*k*_ infects a susceptible individual in *G*_j_, so that either *A*_j_ increases by one (with probability *ξ*_j_) or *P*_j_ increases by one (with probability (1 − *ξ*_j_). This occurs with probability

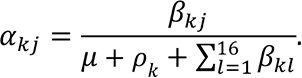
2. The infected individual in *G*_*k*_ recovers, dies or is isolated before infecting anyone else, so that *S*_*k*_ decreases to zero (and there are no infected individuals left in the population). This occurs with probability

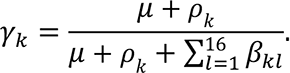

If there are no infectious hosts present in the population, then a local outbreak will not occur. Therefore, assuming that chains of transmission arising from infectious individuals are independent, the probability that no local outbreak occurs beginning from a single symptomatic individual in *G*_*k*_ is

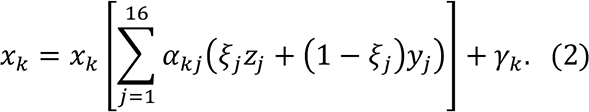

Similarly, beginning instead with a single presymptomatic individual in *G*_*k*_, the possibilities for the next event are:

1. The presymptomatic infected individual in *G*_*k*_ infects a susceptible individual in *G*_j_, so that (as before) either *A*_j_ increases by one (with probability *ξ*_j_) or *P*_j_ increases by one (with probability (1 − *ξ*_j_)). This occurs with probability

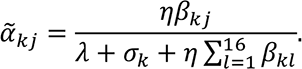
2. The infected individual in *G*_*k*_ develops symptoms (transitions from *P*_*k*_ to *S*_*k*_). This occurs with probability

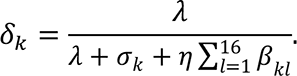
3. The infected individual in *G*_*k*_ is isolated before infecting anyone else, so that *S*_*k*_ decreases by one. This occurs with probability

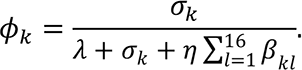

Therefore, the probability that no local outbreak occurs beginning from a single presymptomatic individual in *G*_*k*_ is

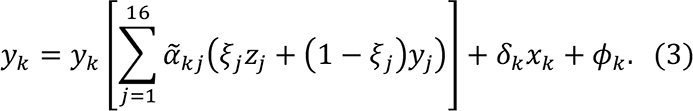

Similarly, the probability *z*_*k*_ that a local outbreak does not occur starting from a single asymptomatic individual in *G*_*k*_ satisfies the equation

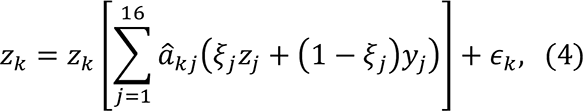

Where

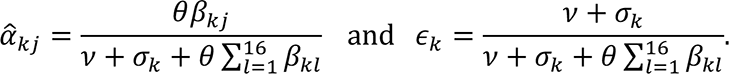

The system of simultaneous equations (2) − (4) can be solved numerically to obtain *x*_*k*_, *x*_*k*_ and *z*_*k*_(here, we did this using the MATLAB nonlinear system solver *‘fsolve’*). Specifically, we take the minimal non-negative solution, as is standard when calculating extinction probabilities using branching process models [55, 68]. Then, for each *k*, the probability of a local outbreak occurring beginning from a single symptomatic (or presymptomatic, asymptomatic respectively) individual in group *G*_*k*_ is given by 1 − *x*_*k*_ (1 − *x*_*k*_, 1 − *z*_*k*_).

Throughout, we consider the probability *p*_*k*_ of a local outbreak occurring beginning from a single nonsymptomatic individual in group *G*_*k*_ arriving in the population at the beginning of their infection:

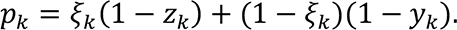

The average local outbreak probability, *P*, which is defined as the probability of a local outbreak when the index case is chosen randomly from the population, is also considered. The value of *P* is therefore a weighted average of the *p*_*k*_ values, where the weights correspond to the proportion of the population represented by each group:

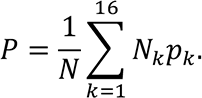

This reflects an assumption that the index case is more likely to come from an age group with more individuals than an age group with fewer individuals.

All computing code used to implement the above methods was written in MATLAB version R2019a, and is available at https://github.com/francescalovellread/age-dependent-outbreak-risks.

## 3. Results

### 3.1 Effect of the age of the index case on the risk of a local outbreak

We first considered the probability that a single infected individual in a particular age group *G*_*k*_ initiates a local outbreak when introduced into a new host population. This quantity was calculated for each of the three scenarios A, B and C (Fig 3).

**Fig 3.**
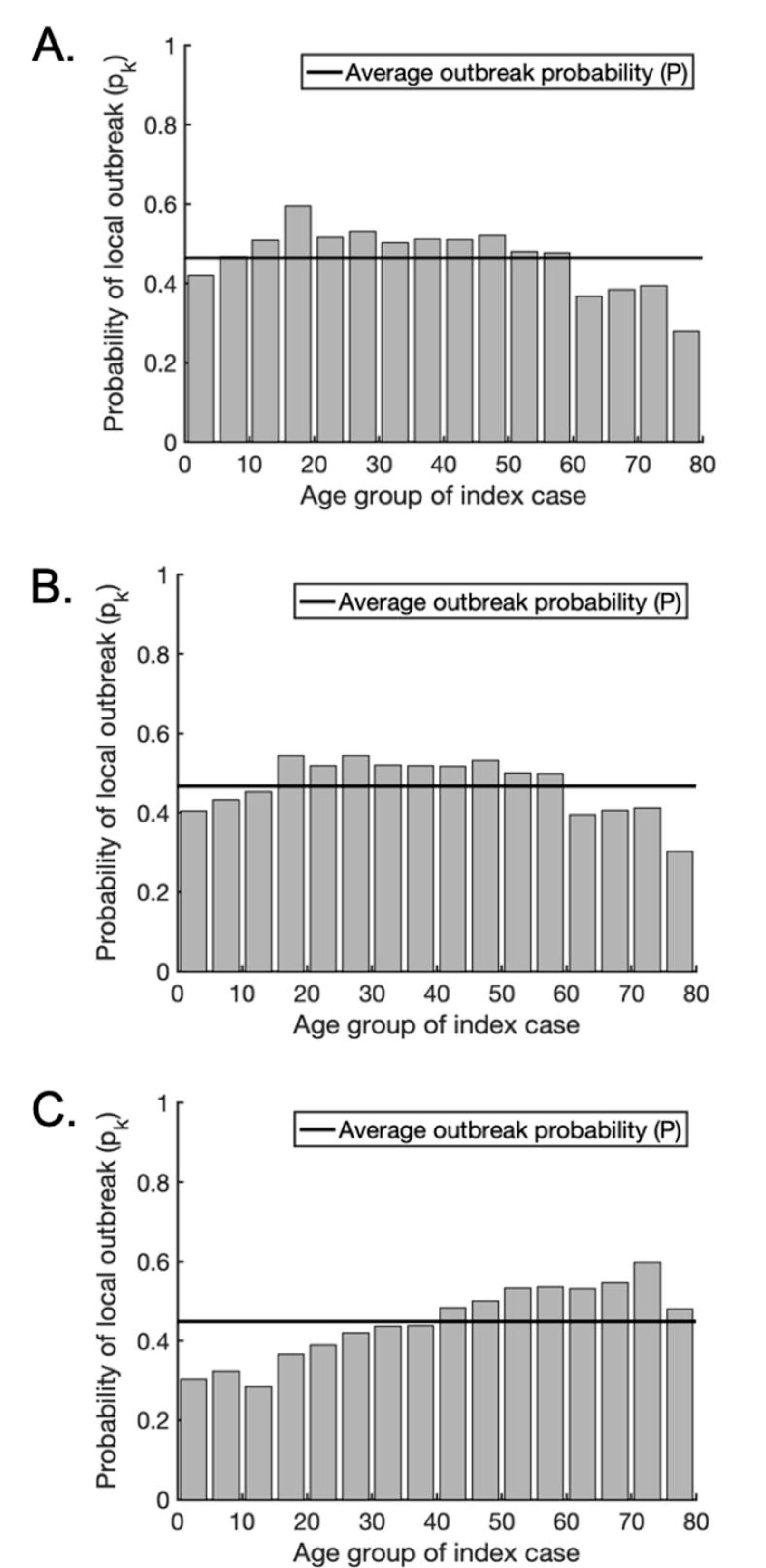
The probability of a local outbreak depends on the age of the index case. A. The probability that a single infected individual in any given age group triggers a local outbreak (grey bars) for scenario A, in which susceptibility and clinical fraction are assumed constant across all age groups. The weighted average local outbreak probability *P* is shown by the black horizontal line. B. The analogous figure to A but for scenario B, in which clinical fraction is assumed constant across all age groups but susceptibility varies with age (Table 1). C. The analogous figure to A but for scenario C, in which both susceptibility and clinical fraction vary with age (Table 1).

In scenario A, the variation in the local outbreak risk for introduced cases of different ages is driven solely by the numbers of contacts between individuals. As a result, due to their higher numbers of daily contacts, school- and working-age individuals are more likely to trigger a local outbreak than children under five or adults over 60, with index cases aged 15-19 posing the highest risk (Fig 3A). These findings do not change significantly when susceptibility is allowed to vary with age in scenario B (Fig 3B). However, in scenario C, assuming that the clinical fraction also varies between age groups alters the age-dependent risk profile substantially. This is because asymptomatic individuals are assumed to be less infectious than symptomatic individuals, and therefore an index case in an age group with a high proportion of asymptomatic infections is less likely to initiate a local outbreak. In this scenario, index cases aged 40 or over had a disproportionately high probability of generating a local outbreak, with individuals aged 70-74 presenting the highest risk (Fig 3C). These individuals are more likely to develop symptoms than younger individuals (Table 1), leading to a higher expected infectiousness. In contrast, individuals under the age of 40 had a below average probability of generating a local outbreak, with individuals aged 10-14 presenting the lowest risk. Noticeably, individuals aged 5- 19 presented relatively low risks, despite the high numbers of contacts occurring among these age groups (Fig 2B). In this scenario, the large number of contacts was offset by the fact that individuals in these age groups are more likely to be asymptomatic and consequently less infectious than older individuals (Table 1). Therefore, an index case in one of these age groups is likely to lead to fewer secondary transmissions. Furthermore, the contact patterns between individuals in these age groups are highly assortative with respect to age (Fig 2B). Therefore, in addition to the index case being less infectious, a high proportion of the contacts they make are with individuals who are also likely to be less infectious, as well as being less susceptible to infection in the first place.

We performed our subsequent analyses for each of the three scenarios A, B and C, with qualitatively similar results. The figures shown in the main text are for scenario C, since we deem this scenario to be the most realistic for SARS-CoV-2 transmission, but the analogous results for scenarios A and B are presented in Supplementary Figs S1-6.

### 3.2 Effect of the target age group on NPI effectiveness

We next considered the effects of NPIs that reduce the number of contacts between individuals on the probability that an introduced case will lead to a local outbreak. To approximate the relative effects of school closures, workplace closures and population-wide social distancing policies, we calculated the age-dependent risk profiles when each of these types of contact were excluded from the overall contact matrix.

First, we removed all ‘school’ contacts from the total contact matrix (Fig 4A). For scenario C, removing ‘school’ contacts led to a 4.2% reduction in the average probability of a local outbreak (from 0.449 to 0.430). This small reduction is unsurprising for scenario C, since in that scenario school-aged infected individuals are assumed to be more likely to be asymptomatic than other infected individuals, and therefore their expected infectiousness is lower. However, even for scenarios A and B, in which school-aged individuals present the greatest risk of triggering a local outbreak, the effectiveness of removing ‘school’ contacts alone at reducing the local outbreak probability was limited (reductions of 7.2% and 4.75% respectively; see Supplementary Figs S1A, S4A). In each scenario, the reduction in risk was predominantly for school-aged index cases, with the risk from index cases of other ages only slightly reduced. Second, we considered the effects of removing ‘work’ contacts from the total contact matrix (Fig 4B). This led to a more substantial 25.4% reduction in the average probability of a local outbreak for scenario C (with corresponding reductions of 19.0% and 24.0% for scenarios A and B respectively; see Supplementary Figs S1B, S4B). As well as reducing the risk of a local outbreak from an index case of working age, removing ‘work’ contacts also reduced the probability of a local outbreak occurring starting from a school-aged individual. This is because closing workplaces helps to block chains of transmission that begin with an infected child. For example, a transmission chain involving a child transmitting to an adult at home, followed by subsequent spread around the adult’s workplace, will be less likely to occur. Third, we investigated the effect of removing all ‘other’ contacts, reflecting perfect social distancing being observed outside of the home, school or workplace (Fig 4C). This had the most significant effect of the three types of contact-reducing intervention considered, reducing the probability of a local outbreak by 41.7% for scenario C (and 30.7% or 33.2% for scenarios A and B, respectively).

**Fig 4.**
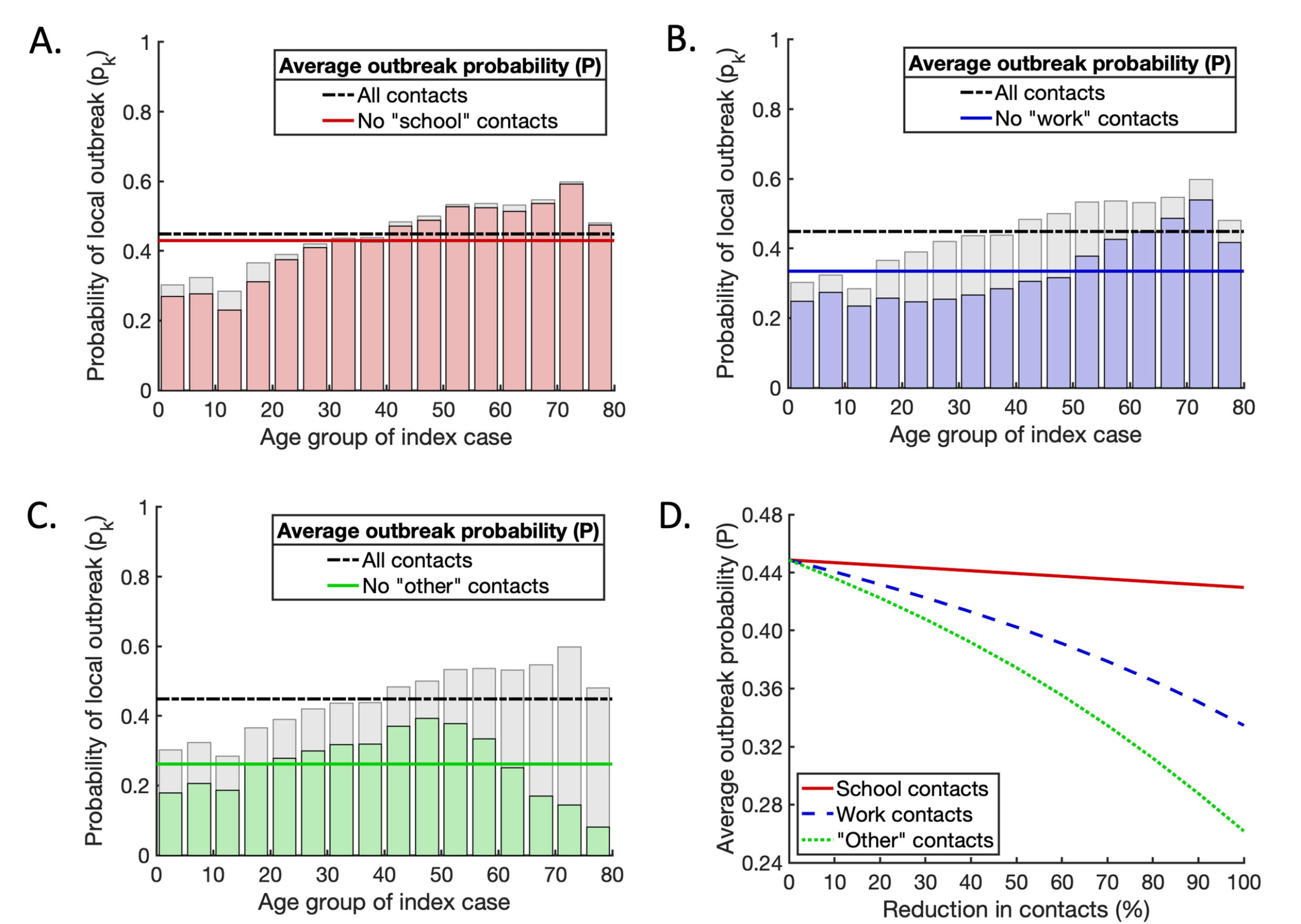
The effects of interventions that reduce contacts between individuals on the probability of a local outbreak. A. The effect of removing all ‘school’ contacts on the probability of a local outbreak. Pale grey bars and black dash-dotted line represent the local outbreak probabilities without any contacts removed (as in Fig 3C). Red bars and the solid red line represent the local outbreak probabilities and their weighted average when ‘school’ contacts are removed. B. The analogous figure to A, but with all ‘work’ contacts removed. C. The analogous figure to A, but with all ‘other’ contacts removed. D. Partial reductions in ‘school’, ‘work’ and ‘other’ contacts, and the resulting reductions in the average local outbreak probability (solid red, dashed blue and dotted green lines respectively).

In the three cases described above, we considered complete reductions in ‘school’, ‘work’ and ‘other’ contacts, respectively. In practice, such complete elimination of contacts is unlikely. We therefore also considered partial reductions in ‘school’, ‘work’ and ‘other’ contacts, and compared the resulting reductions in the local outbreak probability (Fig 4D). For any given percentage reduction in contacts, reducing ‘other’ contacts always led to the largest reduction in the local outbreak probability (see also Supplementary Figs S1D, S4D). This suggests that reducing social contacts outside schools and workplaces can be an important component of strategies to reduce the risk of local outbreaks of SARS-CoV-2. However, this alone is not enough to eliminate the risk of local outbreaks entirely. For greater risk reductions using contact-reducing NPIs, a mixed approach involving combinations of reductions in ‘school’, ‘workplace’ and ‘other’ contacts is needed.

### 3.3 Mixed strategies for reducing the local outbreak risk

Next, we considered the effects of combining reductions in ‘school’, ‘work’ and ‘other’ contacts on the local outbreak probability (Fig 5; analogous results for scenarios A and B are shown in Supplementary Figs S2 and S5). We allowed reductions in ‘school’ and ‘work’ contacts to vary between 0% and 100% whilst ‘other’ contacts were reduced by 25%, 50% or 75% (Fig 5A,B,C, respectively).

**Fig 5.**
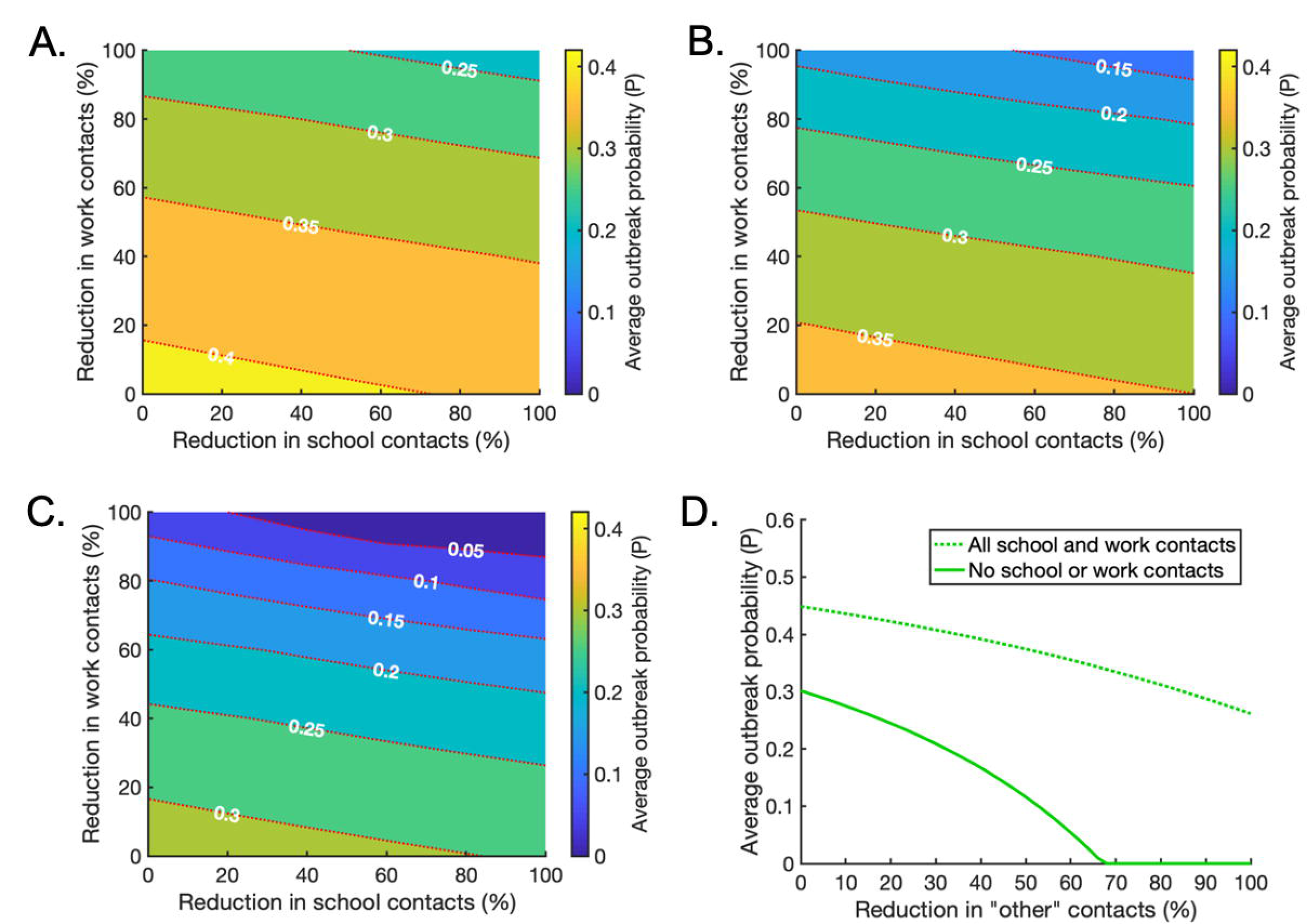
The effects of intervention strategies that combine reductions in ‘school’, ‘work’ and ‘other’ contacts. A. The effect of reducing ‘school’ and ‘work’ contacts on the weighted average probability of a local outbreak (*P*), when ‘other’ contacts are reduced by 25% across all age groups. Red dotted lines indicate contours along which the local outbreak probability is constant. B. The analogous figure to A, but with a 50% reduction in ‘other’ contacts. C. The analogous figure to A, but with a 75% reduction in ‘other’ contacts. D. The effect of reducing ‘other’ contacts on the average local outbreak probability when ‘school’ and ‘work’ contacts are not reduced at all (dotted line) and when ‘school’ and ‘work’ contacts are reduced by 100% (solid line).

Since NPIs have negative economic, social and non-disease health consequences, policy makers may choose to implement public health measures in which the risk of local outbreaks is not eliminated completely. These results provide contact reduction targets for mixed strategies in which the local outbreak probability is reduced to a pre-specified ‘acceptable’ level. For example, to reduce the local outbreak probability to 0.25, ‘other’ contacts could be reduced by 25% from the baseline level, and ‘school’ and ‘home’ contacts reduced as indicated by the red dotted contour marked ‘0.25’ in Fig 5A. Alternatively, to achieve the same local outbreak risk, ‘other’ contacts can instead be reduced by 50% or by 75% with the degree of ‘school’ and ‘work’ reductions chosen according to the contours marked ‘0.25’ in Figs 5B,C respectively.

If a policy maker wishes to eliminate the local outbreak risk entirely using contact-reducing NPIs, for the model parameterisation considered very significant reductions in multiple types of contacts are needed in combination. For example, even if all ‘school’ and ‘work’ contacts are removed, ‘other’ contacts must be reduced by 66% for the overall average local outbreak probability to fall below 0.01 (Fig 5D). Since such substantial reductions in multiple types of contacts are unlikely to be possible, this suggests that contact-reducing NPIs must be combined with other interventions, such as effective surveillance and isolation strategies, to eliminate local outbreaks.

### 3.4 Effect of surveillance on the contact-reducing NPIs required for local outbreak control

We considered whether or not low local outbreak probabilities can be achieved using limited contact-reducing NPIs in combination with other interventions. Specifically, the effects of surveillance and isolation of infected individuals (through e.g. contact tracing) as well as reducing contacts in schools, workplaces and other locations, were assessed. While results are shown for scenario C in Fig 6, analogous results for scenarios A and B are presented in Supplementary Figs S3 and S6.

**Fig 6.**
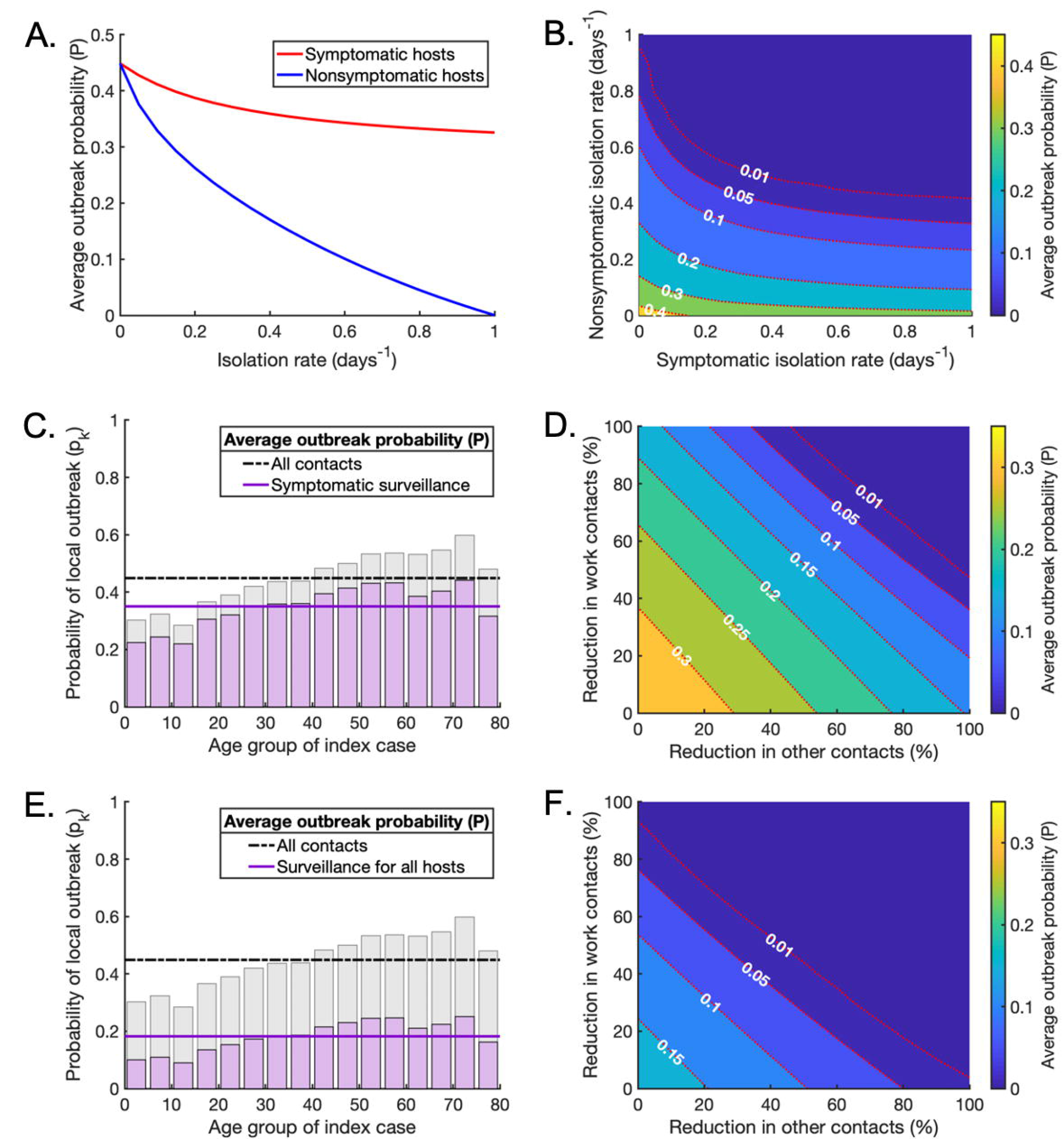
Surveillance as part of a mixed strategy to reduce the local outbreak probability. A. The effect of increasing the isolation rate of symptomatic (red line) or nonsymptomatic infected hosts (blue line) on the average probability of a local outbreak (*P*), in the absence of contact-reducing NPIs. The isolation rates *ρ*_*k*_ and *σ*_*k*_ are varied in turn between 0 days^’1^ and 1 days^’1^. B. The effect of simultaneously varying the isolation rate of symptomatic and nonsymptomatic hosts on the average probability of a local outbreak (*P*), again without contact-reducing NPIs. C. The age-dependent probability of a local outbreak when the isolation rate of symptomatic individuals is *ρ*_*k*_ = 1/2 days^’1^, without contact-reducing NPIs or surveillance of nonsymptomatic infected individuals (purple bars and solid line). Pale grey bars and black dash-dotted line represent the original local outbreak probabilities without any contact-reducing NPIs or enhanced surveillance (as in Fig 3C). D. The effect of reducing ‘work’ and ‘other’ contacts when the isolation rate of symptomatic infected individuals is *ρ*_*k*_ = 1/2 days^’1^, as in C, without surveillance of nonsymptomatic infected individuals. E,F. The analogous figures to C,D, with enhanced surveillance of both symptomatic and nonsymptomatic infected hosts (*ρ*_*k*_ = 1/2 days^’1^ and *σ*_*k*_ = 1/7 days^’1^).

Initially, we considered the effect of increasing the rate at which symptomatic and/or nonsymptomatic infected individuals are detected and isolated as a result of surveillance, in the absence of contact-reducing NPIs (i.e. with no reduction in the number of contacts between individuals compared to the baseline case in Fig 2B) (Fig 6A,B). For symptomatic hosts, this represents an enhanced rate of isolation compared to the baseline rate of self-isolation already present in the model. Isolation of nonsymptomatic hosts was more effective at reducing the local outbreak probability than isolation of symptomatic hosts (Figs 6A,B), although of course this is more challenging to achieve [55]. However, if fast isolation of nonsymptomatic hosts could be achieved through efficient large-scale testing (potentially in combination with contact tracing [69]), the probability of local outbreaks could be reduced substantially through this measure alone.

We then demonstrated the effects of combining contact-reducing NPIs with enhanced isolation of infected hosts due to infection surveillance. First, we increased the enhanced isolation rate of symptomatic individuals to *ρ*_*k*_ = 1/2 days^)1^. In the absence of other interventions, this reduced the local outbreak probability by 22.0% (Figure 6C). With this level of surveillance, the local outbreak risk could be reduced below 0.01 with a reduction in ‘work’ and ‘other’ contacts of around 73% each, for example.

Finally, keeping the enhanced isolation rate of symptomatic individuals equal to *ρ*_*k*_ = 1/2 days^)1^, we increased the isolation rate of nonsymptomatic individuals to *σ*_*k*_ = 1/7 days^)1^. In this case, the local outbreak probability without contact-reducing NPIs fell by 59.4% compared to a situation without enhanced surveillance (Fig 6E), and the reductions in ‘work’ and ‘other’ contacts needed to bring the local outbreak probability below 0.01 were significantly smaller (Figure 6F). For example, if ‘work’ contacts can be reduced by 50%, then ‘other’ contacts only need to be reduced by 43%. This indicates that effective surveillance of both symptomatic and nonsymptomatic individuals can substantially lower the extent of contact- reducing NPIs that are required to achieve substantial reductions in local outbreak risks.

## 4. Discussion

During the COVID-19 pandemic, public health measures that reduce the numbers of contacts between individuals have been implemented in countries globally. These measures include school closures, workplace closures and population-wide social distancing policies. Contact-reducing NPIs have been shown to be effective at reducing SARS-CoV-2 transmission, and have also been used previously during influenza pandemics [5, 70–72]. However, long-term implementation of these measures has negative social, psychological and economic consequences [7, 12–16]. It is therefore important to assess the effectiveness of different contact-reducing NPIs at lowering transmission and preventing local outbreaks, in order to design effective targeted control strategies that avoid unnecessarily strict measures.

Here, we constructed a branching process model to estimate the risk of local outbreaks under different contact-reducing NPIs and different levels of surveillance for symptomatic and nonsymptomatic infected individuals. Unlike previous approaches for estimating outbreak risks using branching processes [51–56], we considered the effects of age-related heterogeneities affecting transmission for infected individuals of a wide range of ages, including age-dependent variations in social mixing patterns, susceptibility to infection and clinical fraction. Using SARS-CoV-2 as a case study, we demonstrated that the risk that an introduced case initiates a local outbreak depends on these age-related factors and on the age of the introduced case (Fig 3), as well as the age-structure of the local population.

We used our model to assess the effects of reducing the numbers of contacts that occur in school, in the workplace and elsewhere. Of the three contact-reducing NPIs considered, removing ‘school’ contacts had the smallest effect on the probability of observing a local outbreak, even when age-dependent variations in susceptibility and clinical fraction were ignored (Figs 4A,D, Supplementary Figs S1A,D, and S4,A,D). This can be attributed to the fact that school closures predominantly reduce contacts between individuals aged 5-19, who only account for approximately 23% of the total population [58]. Additionally, these individuals tend to have large numbers of contacts outside of the school environment (Figs 2E,F). Therefore, compared to other measures, interrupting within-school transmission may have only a limited effect on transmission in the wider population, particularly when school-aged individuals are less susceptible to infection and more likely to experience subclinical courses of infection. In contrast, reducing contacts that occur outside schools or workplaces was the most effective intervention, significantly lowering the local outbreak risk across all age groups, and for those aged over 60 in particular (Figs 4C,D, and Supplementary Figs S1C,D and S4C,D). This could explain the success of social distancing strategies worldwide for reducing observed COVID-19 cases and deaths.

Mixed strategies combining reductions in ‘school’, ‘work’ and ‘other’ contacts led to greater reductions in the local outbreak probability than individual interventions (Figs 5A-C), but very large reductions in all three types of contact were required to eliminate the risk of local outbreaks entirely (Fig 5D). However, implementing effective surveillance to identify infected hosts (followed by isolation) led to substantial reductions in the risk of local outbreaks even in the absence of other control measures (Figs 6A,B). In the scenarios considered here, with an efficient surveillance strategy in place, significantly smaller reductions in ‘work’ and ‘other’ contacts were needed to render the local outbreak probability negligible, even when ‘school’ contacts were not reduced at all (Figs 6C-F). This supports the use of surveillance that targets both symptomatic and nonsymptomatic individuals, such as contact tracing and isolation strategies or population-wide diagnostic testing, to help prevent local outbreaks [55].

Although here we used SARS-CoV-2 as a case study, our model provides a framework for estimating the risk of local outbreaks in age-structured populations that can be adapted for other pathogens, provided sufficient data are available to parametrise the model appropriately. The effects of age-structure on local outbreak risks may vary for pathogens with different epidemiological characteristics. For influenza-A viruses, for example, susceptibility to infection tends to decrease with age, whilst the risk of an infection leading to severe symptoms is greater both for the elderly and for the very young [31, 73, 74]. This is in contrast to SARS-CoV-2, for which children are more likely to experience subclinical courses of infection. In this study, we used age demographic and contact data for the UK, but equivalent data for other countries are available and can easily be substituted into our model to estimate outbreak risks elsewhere [17, 58].

One caveat of the results for SARS-CoV-2 presented here is that, although the epidemiological parameters of our model were chosen to be consistent with reported literature estimates, there is considerable variation between studies. In particular, the precise age-dependent variation in susceptibility and clinical fraction remains unclear, and the relative infectiousness of asymptomatic, presymptomatic and symptomatic hosts has not been determined exactly.

Furthermore, the inherent transmissibility of SARS-CoV-2 is now higher than in the initial stage of the pandemic, due to the appearance of novel variants. To explore ongoing local outbreak risks due to SARS-CoV-2, it would be necessary to update the model to reflect the increased transmissibility of the Delta variant [75]. Due to the uncertainty in model parameter values, we conducted sensitivity analyses to explore the effects of varying the parameters of the model on our results (Supplementary Figs S1-12). In each case that we considered, our main conclusions were unchanged: the probability that an introduced case initiates a local outbreak depends on age-dependent factors affecting pathogen transmission and control, with widespread interventions and combinations of NPIs reducing the risk of local outbreaks most significantly.

An important limitation of our approach to modelling contact-reducing NPIs is that we made a standard assumption in our main analyses that ‘school’, ‘work’ and ‘other’ contacts are independent [27, 31, 47]. In other words, reducing the numbers of contacts in one location did not affect the numbers of contacts occurring in another. In reality, this is unlikely to be the case. For example, closing schools also affects workplace contacts, as adults may then work from home in order to fulfil childcare requirements. Additionally, the contact data that we used represent the number of unique contacts per day and do not reflect the numbers of repeated contacts with the same person, which affect the risk of transmission between individuals. These assumptions could in principle be removed, if relevant data become available – for example, data describing the effects of school closures on numbers of contacts in other locations. To demonstrate how changes in multiple types of contact due to NPIs could be implemented in the model, we conducted a supplementary analysis in which we investigated the effects of removing all ‘school’ contacts and allowing for concurrent changes in ‘work’, ‘home’ and ‘other’ contacts (Supplementary Figs S13-14). These results support our conclusion that school closures are unlikely to have a substantial impact on SARS-CoV-2 transmission when applied as the sole NPI. The benefit of school closures could even potentially be outweighed by the possible secondary effects on other types of contacts. An improved understanding of how NPIs affect different types of contact is important for more accurate assessments of interventions in future.

Despite the simplifications made, our model provides a useful framework for estimating the risk of local outbreaks and the effects of NPIs. Different measures can be considered in combination in the model to develop strategies for lowering local outbreak risks. Our results emphasise the importance of quantifying age-dependent factors that affect transmission dynamics, such as susceptibility to infection and the proportion of hosts who develop clinical symptoms, for individuals of different ages. As we have shown, it is crucial to take age-dependent factors into account when assessing local outbreak risks and designing public health measures.

## Authors’ contributions

Conceptualisation: All authors. Methodology: FALR, RNT. Investigation: FALR, SS. Writing - original draft: FALR, RNT. Writing - review and editing: All authors. Supervision: RNT.

## Data availability

All computer code used in this paper was written in MATLAB version R2019a, and is available at the following GitHub repository: https://github.com/francescalovellread/age-dependent-outbreak-risks.

