## Supplementary Material for "Estimating local outbreak risks and the effects of non-pharmaceutical interventions in age-structured populations: SARS-CoV-2 as a case study"

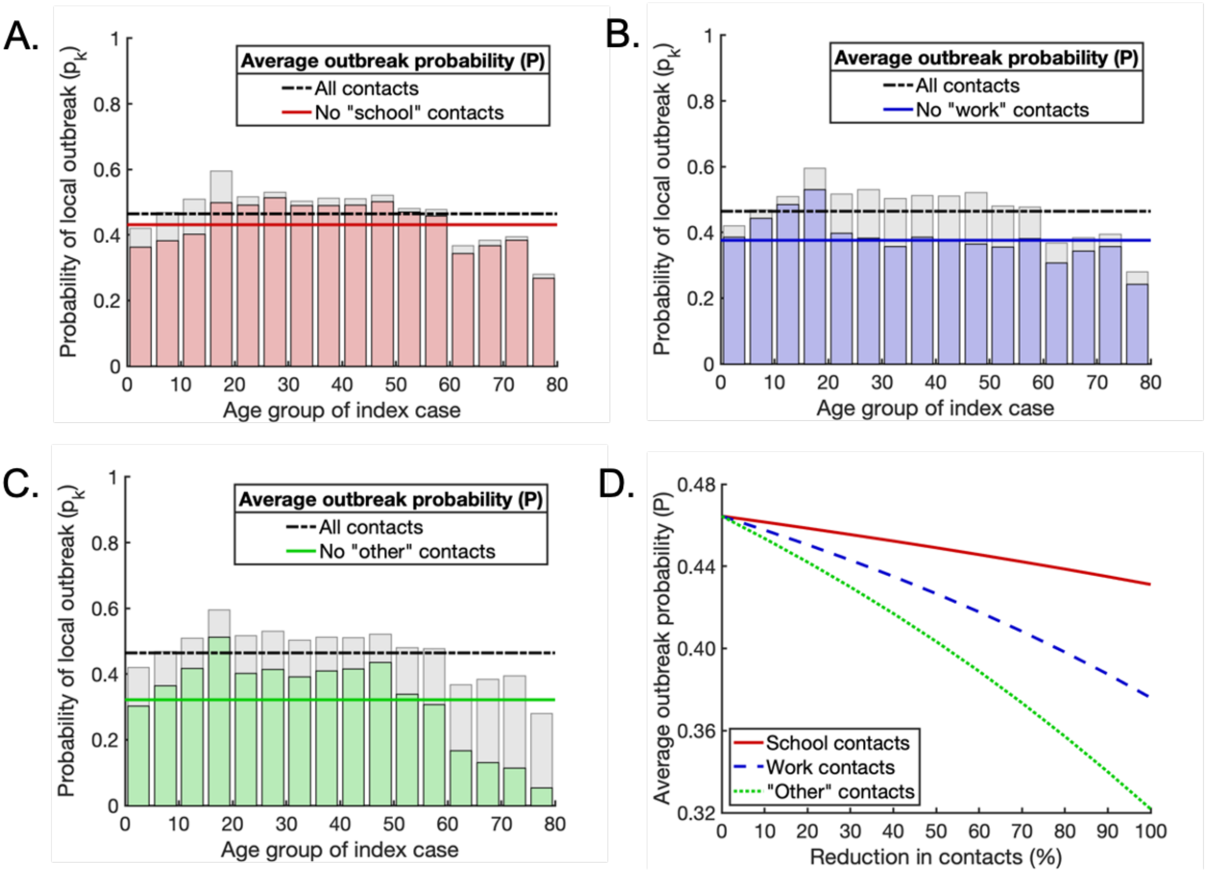


**Fig S1. Scenario A: The effects of interventions that reduce contacts between individuals on the probability of a local outbreak.** For scenario A, susceptibility to infection and the proportion of hosts who experience a fully asymptomatic course of infection are independent of age. A. The effect of removing all 'school' contacts on the probability of a local outbreak. Pale grey bars and black dash-dotted line represent the local outbreak probabilities without any contacts removed (as in Fig 3A). Red bars and the solid red line represent the local outbreak probabilities and their weighted average when 'school' contacts are removed. B. The analogous figure to A, but with all 'work' contacts removed. C. The analogous figure to A, but with all 'other' contacts removed. D. Partial reductions in 'school', ‘work’ and ‘other’ contacts, and the resulting reductions in average local outbreak probability (solid red, dashed blue and dotted green lines respectively).

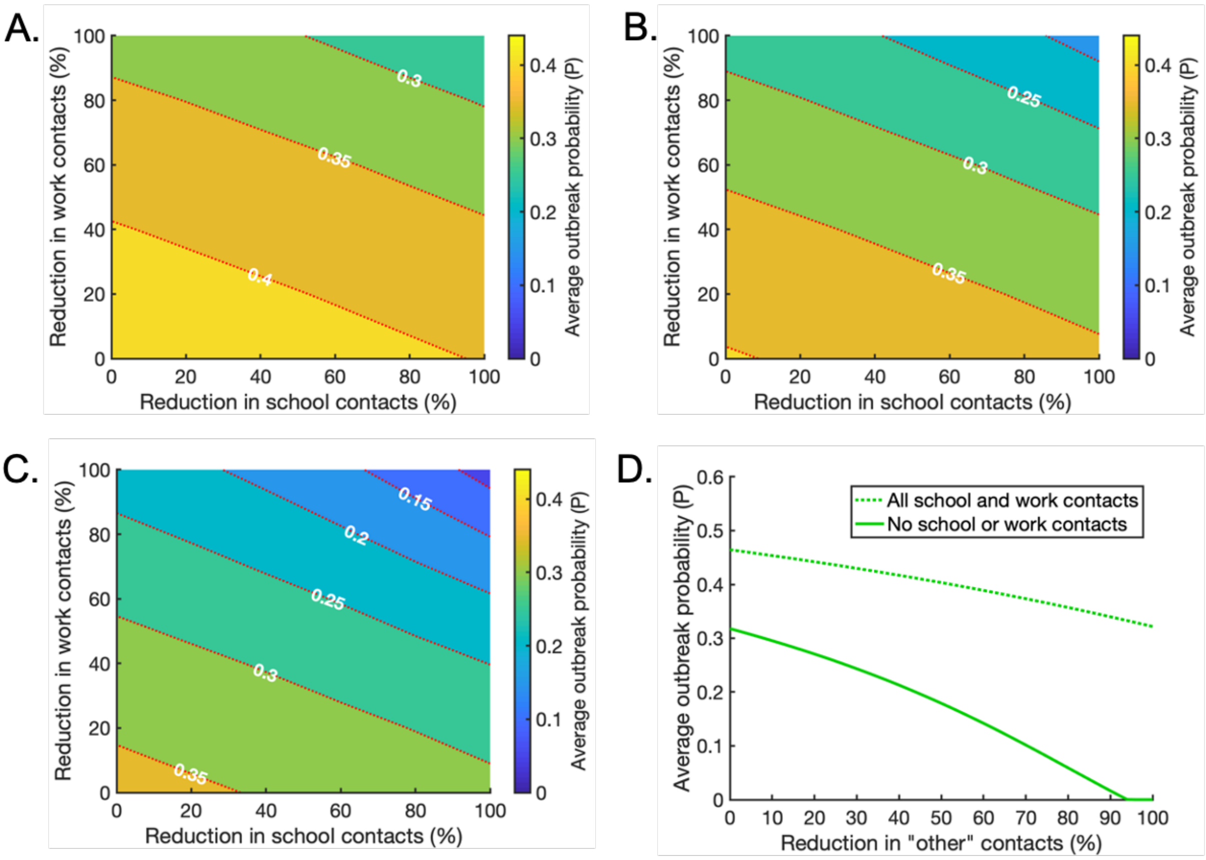


**Fig S2. Scenario A: The effects of intervention strategies that combine reductions in ‘school’, ‘work’ and ‘other’ contacts.** For scenario A, susceptibility to infection and the proportion of hosts who experience a fully asymptomatic course of infection are independent of age. A. The effect of reducing ‘school’ and ‘work’ contacts on the weighted average probability of a local outbreak (*P*), when ‘other’ contacts are reduced by 25% across all age groups. Red dotted lines indicate contours along which the local outbreak probability is constant. B. The analogous figure to A, but with a 50% reduction in ‘other’ contacts. C. The analogous figure to A, but with a 75% reduction in ‘other’ contacts. D. The effect of reducing ‘other’ contacts on the average local outbreak probability when ‘school’ and ‘work’ contacts are not reduced at all (dotted line) and when ‘school’ and ‘work’ contacts are reduced by 100% (solid line).

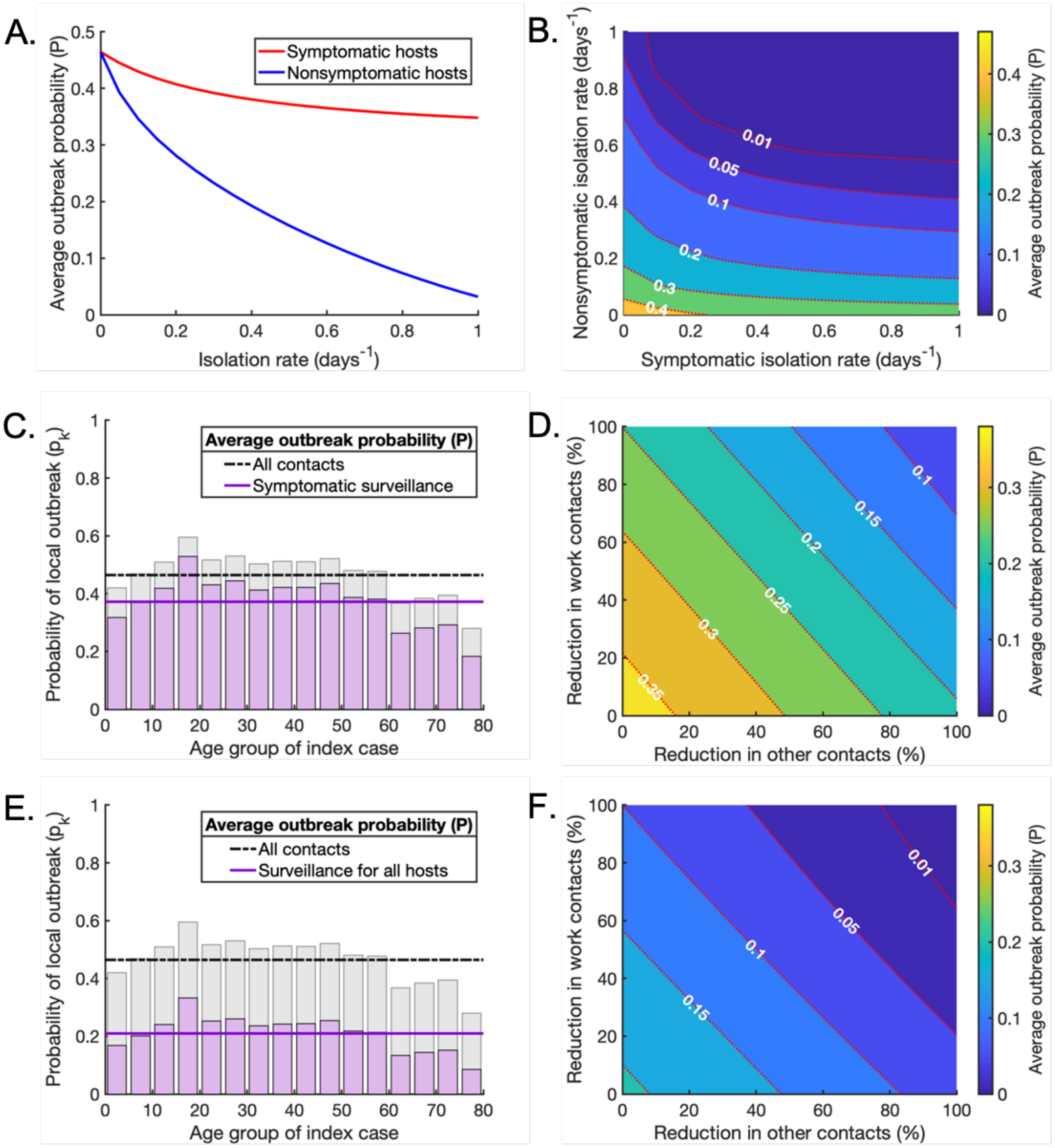


**Fig S3. Scenario A: Surveillance as part of a mixed strategy to reduce the local outbreak probability.** For scenario A, susceptibility to infection and the proportion of hosts who experience a fully asymptomatic course of infection are independent of age. A. The effect of increasing the isolation rate of symptomatic (red line) or nonsymptomatic infected hosts (blue line) on the average probability of a local outbreak ($P$), in the absence of contact-reducing NPIs. The isolation rates $\rho_k$ and $\sigma_k$ are varied between 0 days$^{-1}$ and 1 days$^{-1}$. B. The effect of simultaneously varying the isolation rate of symptomatic and nonsymptomatic hosts on the average probability of a local outbreak ($P$), again without contact-reducing NPIs. C. The age-dependent probability of a local outbreak when the isolation rate for symptomatic individuals is $\rho_k = 1/2 \text{ days}^{-1}$, without contact-reducing NPIs or surveillance of nonsymptomatic infected individuals (purple bars and solid line). Pale grey bars and black dash-dotted line represent the local outbreak probabilities without any contact-reducing NPIs or enhanced surveillance (as in Fig 3A). D. The effect of reducing 'work' and 'other' contacts when the isolation rate of symptomatic individuals is $\rho_k = 1/2 \text{ days}^{-1}$, as in C, without surveillance of nonsymptomatic infected individuals. E,F. The analogous figures to C,D, with enhanced surveillance of both symptomatic and nonsymptomatic infected hosts ($\rho_k = 1/2 \text{ days}^{-1}$ and $\sigma_k = 1/7 \text{ days}^{-1}$).

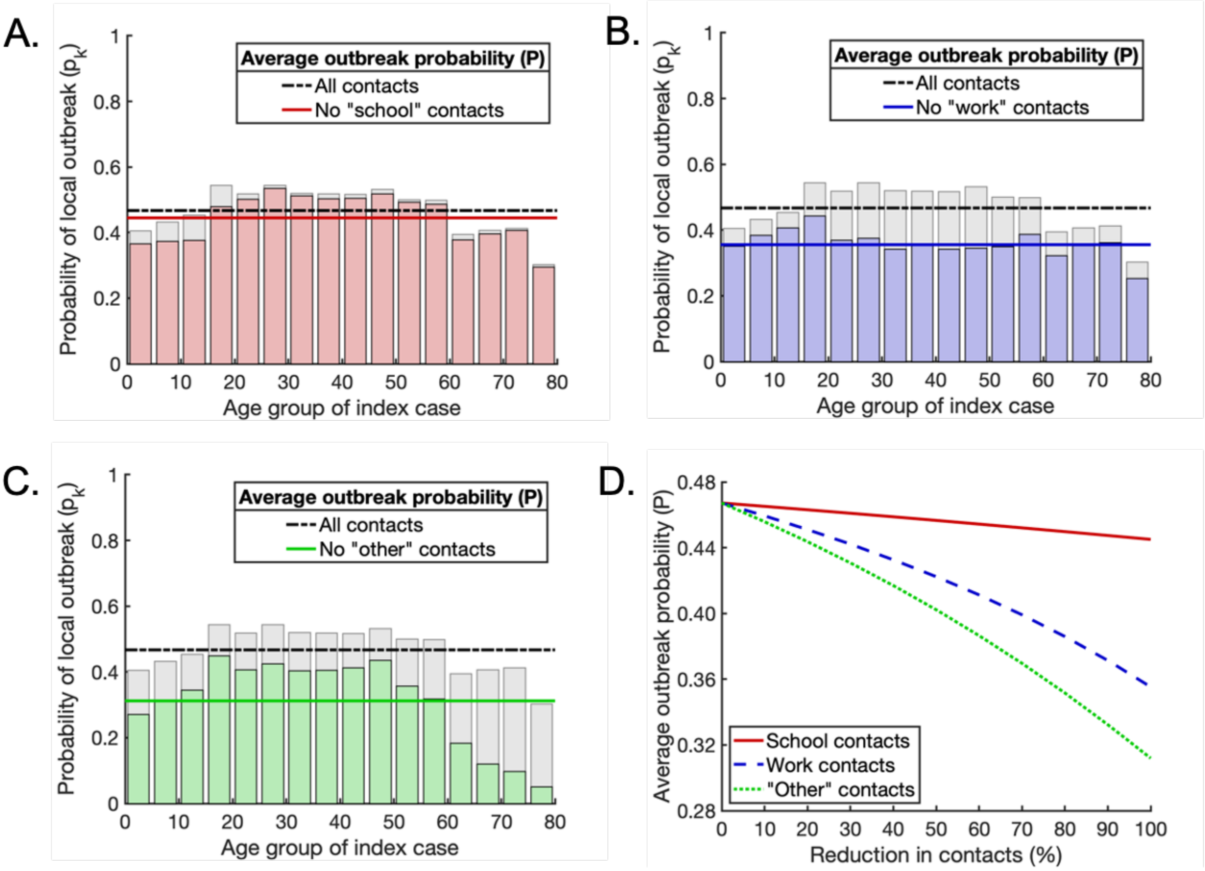


**Fig S4. Scenario B: The effects of interventions that reduce contacts between individuals on the probability of a local outbreak.** For scenario B, susceptibility to infection varies with age but the proportion of hosts who experience a fully asymptomatic course of infection are independent of age. A. The effect of removing all 'school' contacts on the probability of a local outbreak. Pale grey bars and black dash-dotted line represent the local outbreak probabilities without any contacts removed (as in Fig 3B). Red bars and the solid red line represent the local outbreak probabilities and their weighted average when 'school' contacts are removed. B. The analogous figure to A, but with all 'work' contacts removed. C. The analogous figure to A, but with all 'other' contacts removed. D. Partial reductions in 'school', 'work' and 'other' contacts, and the resulting reductions in average local outbreak probability (solid red, dashed blue and dotted green lines respectively).

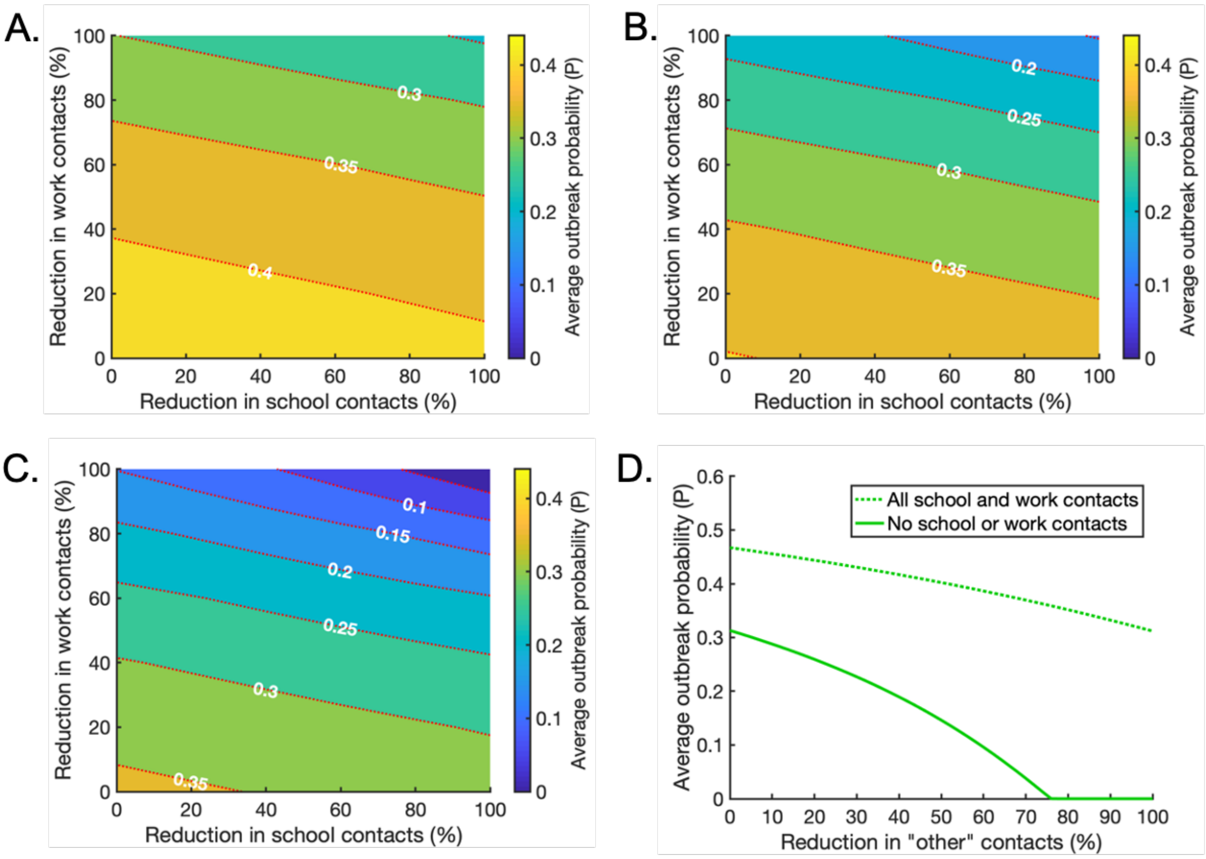


**Fig S5. Scenario B: The effects of intervention strategies that combine reductions in 'school', 'work' and 'other' contacts.** For scenario B, susceptibility to infection varies with age but the proportion of hosts who experience a fully asymptomatic course of infection are independent of age. A. The effect of reducing 'school' and 'work' contacts on the weighted average probability of a local outbreak (*P*), when 'other' contacts are reduced by 25% across all age groups. Red dotted lines indicate contours along which the local outbreak probability is constant. B. The analogous figure to A, but with a 50% reduction in 'other' contacts. C. The analogous figure to A, but with a 75% reduction in 'other' contacts. D. The effect of reducing 'other' contacts on the average local outbreak probability when 'school' and 'work' contacts are not reduced at all (dotted line) and when 'school' and 'work' contacts are reduced by 100% (solid line).

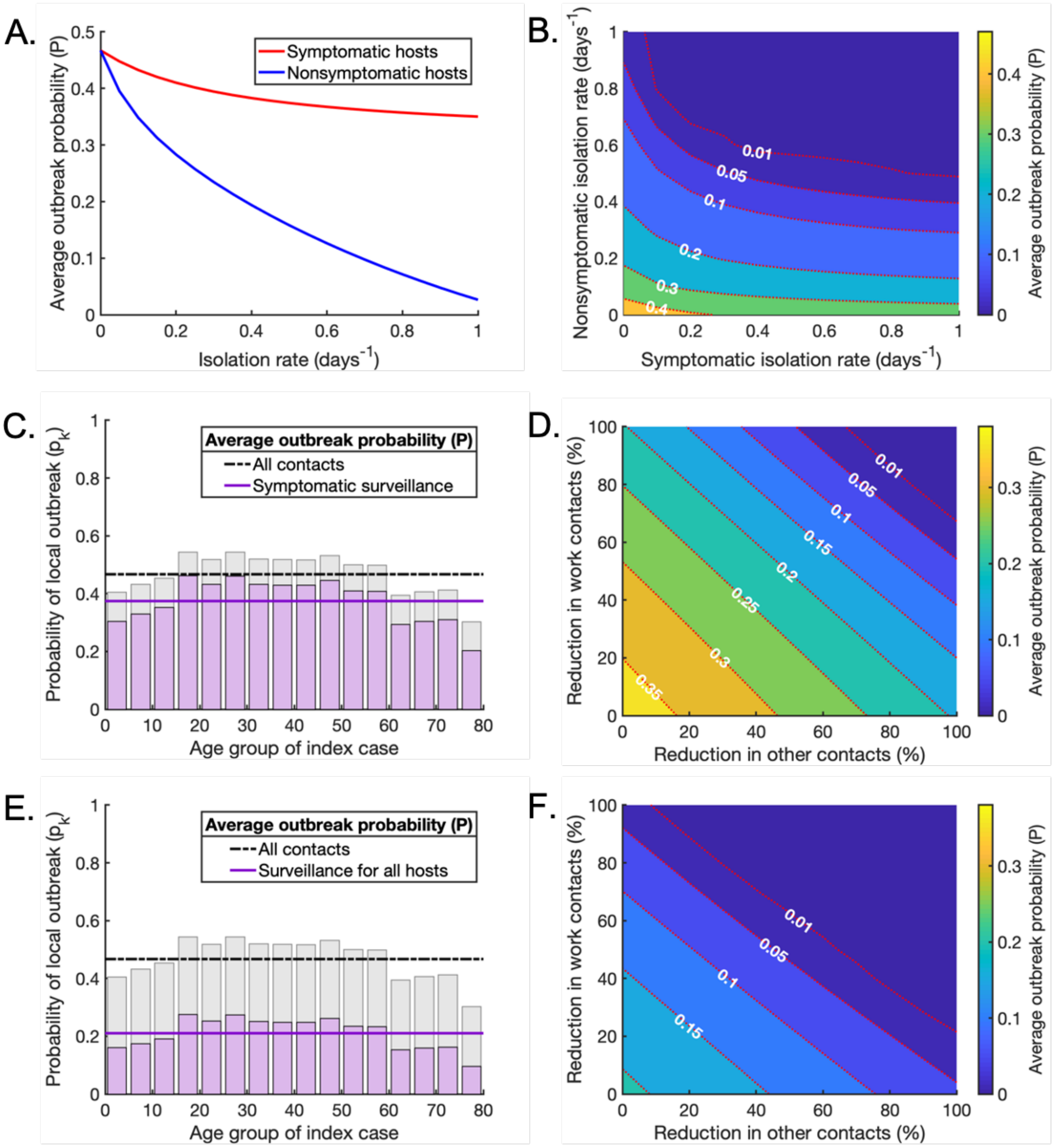


**Fig S6. Scenario B: Surveillance as part of a mixed strategy to reduce the local outbreak probability.** For scenario B, susceptibility to infection varies with age but the proportion of hosts who experience a fully asymptomatic course of infection are independent of age. A. The effect of increasing the isolation rate of symptomatic (red line) or nonsymptomatic infected hosts (blue line) on the average probability of a local outbreak ($P$), in the absence of contact-reducing NPIs. The isolation rates $\rho_k$ and $\sigma_k$ are varied between 0 days$^{-1}$ and 1 days$^{-1}$. B. The effect of simultaneously varying the isolation rate of symptomatic and nonsymptomatic hosts on the average probability of a local outbreak ($P$), again without contact-reducing NPIs. C. The age-dependent probability of a local outbreak when the isolation rate of symptomatic individuals is $\rho_k = 1/2 \text{ days}^{-1}$, without contact-reducing NPIs or surveillance of nonsymptomatic infected individuals (purple bars and solid line). Pale grey bars and black dash-dotted line represent the local outbreak probabilities without any contact reducing NPIs or enhanced surveillance (as in Fig 3B). D. The effect of reducing 'work' and 'other' contacts when the isolation rate of symptomatic individuals is $\rho_k = 1/2 \text{ days}^{-1}$, as in C, without surveillance of nonsymptomatic infected individuals. E,F. The analogous figures to C,D, with enhanced surveillance of both symptomatic and nonsymptomatic infected hosts ($\rho_k = 1/2 \text{ days}^{-1}$ and $\sigma_k = 1/7 \text{ days}^{-1}$).

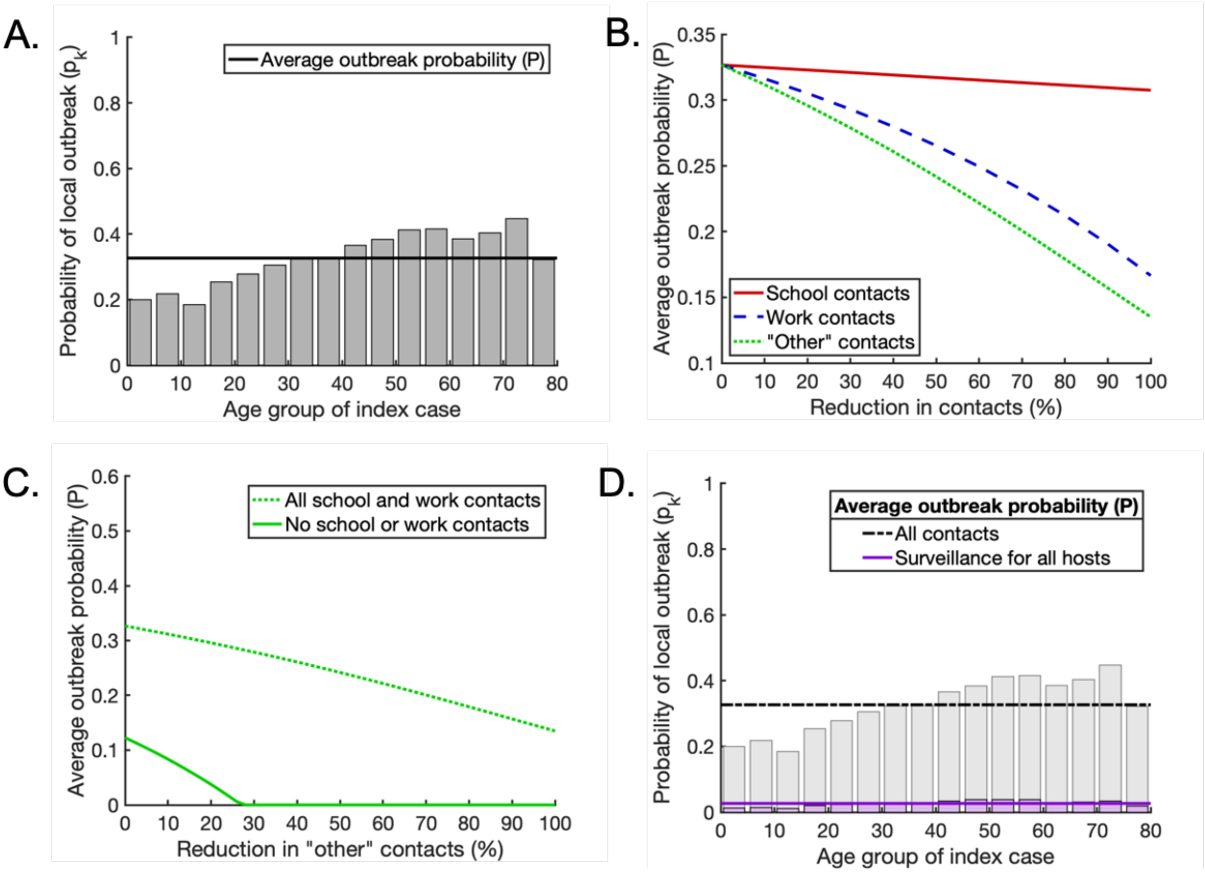


**Fig S7. Scenario C: The effect of reducing the basic reproduction number from $R_0 = 3$ (baseline value) to $R_0 = 2$.** For scenario C, both susceptibility to infection and the proportion of hosts who experience a fully asymptomatic course of infection vary with age. A. Analogous to Fig 3C in the main text: the probability that a single infected individual in any given age group triggers a local outbreak (grey bars) and the weighted average local outbreak probability $P$ (black horizontal line). B. Analogous to Fig 4D in the main text: partial reductions in 'school', 'work' and 'other' contacts, and the resulting reductions in the average local outbreak probability $P$ (solid red, dashed blue and dotted green lines respectively). C. Analogous to Fig 5D in the main text: the effect of reducing 'other' contacts on the average local outbreak probability when 'school' and 'work' contacts are not reduced at all (dotted line) and when 'school' and 'work' contacts are reduced by 100% (solid line). D. Analogous to Fig 6E in the main text: the age-dependent probability of a local outbreak with enhanced surveillance of both symptomatic and nonsymptomatic infected hosts ($\rho_k = 1/2 \text{ days}^{-1}$ and $\sigma_k = 1/7 \text{ days}^{-1}$), without contact-reducing NPIs (purple bars and solid line). Pale grey bars and black dash-dotted line represent the local outbreak probabilities without any contact-reducing NPIs or enhanced surveillance (as in Fig S7A).

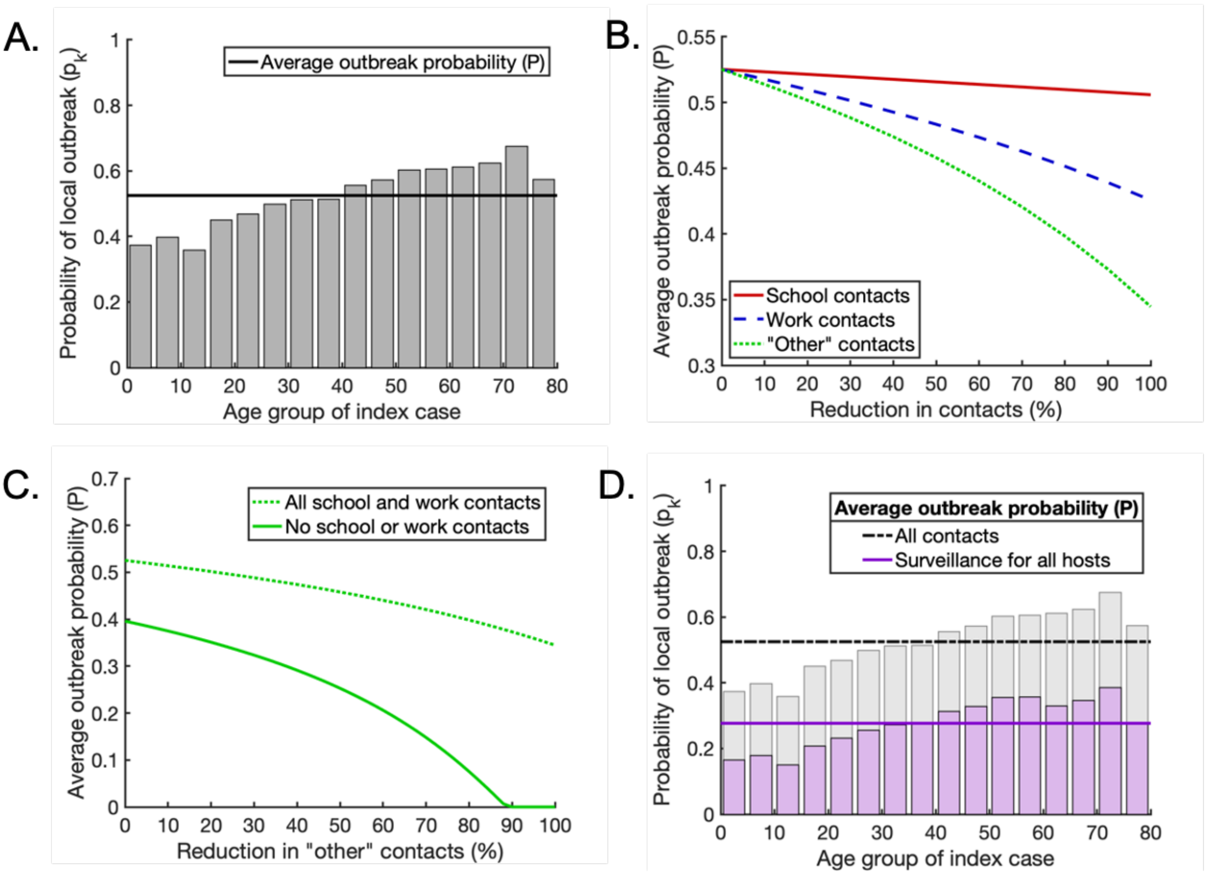


**Fig S8. Scenario C: The effect of increasing the basic reproduction number from $R_0 = 3$ (baseline value) to $R_0 = 4$.** For scenario C, both susceptibility to infection and the proportion of hosts who experience a fully asymptomatic course of infection vary with age. A. Analogous to Fig 3C in the main text: the probability that a single infected individual in any given age group triggers a local outbreak (grey bars) and the weighted average local outbreak probability $P$ (black horizontal line). B. Analogous to Fig 4D in the main text: partial reductions in 'school', 'work' and 'other' contacts, and the resulting reductions in the average local outbreak probability $P$ (solid red, dashed blue and dotted green lines respectively). C. Analogous to Fig 5D in the main text: the effect of reducing 'other' contacts on the average local outbreak probability when 'school' and 'work' contacts are not reduced at all (dotted line) and when 'school' and 'work' contacts are reduced by 100% (solid line). D. Analogous to Fig 6E in the main text: the age-dependent probability of a local outbreak with enhanced surveillance of both symptomatic and nonsymptomatic infected hosts ($\rho_k = 1/2$ days$^{-1}$ and $\sigma_k = 1/7$ days$^{-1}$), without contact-reducing NPIs (purple bars and solid line). Pale grey bars and black dash-dotted line represent the local outbreak probabilities without any contact-reducing NPIs or enhanced surveillance (as in Fig S8A).

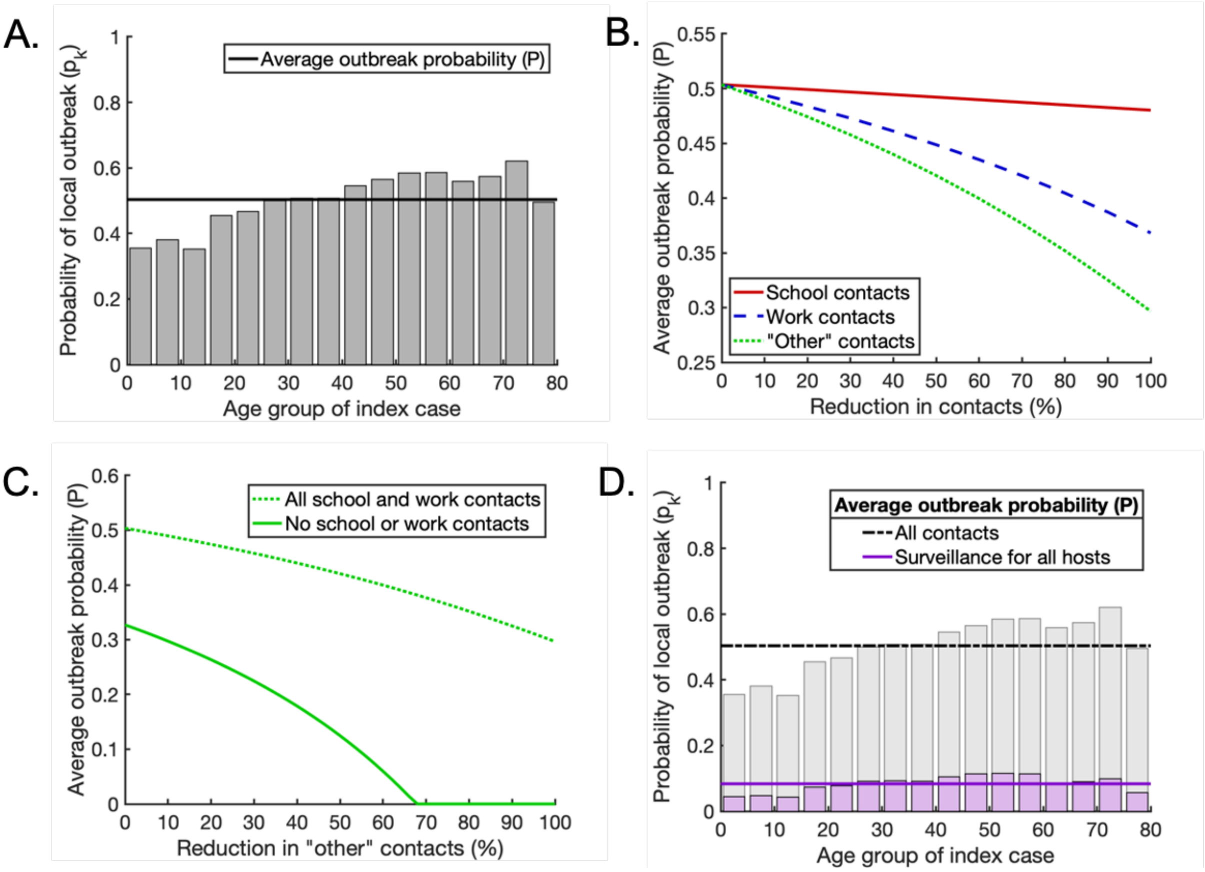

**Fig S9. Scenario C: The effect of reducing the proportion of infections that arise from presymptomatic hosts from $K_p = 0.489$ (baseline value) to $K_p = 0.25$.** The proportions of infections arising from symptomatic and asymptomatic hosts are adjusted so that they remain in the same ratio as in the baseline case. For scenario C, both susceptibility to infection and the proportion of hosts who experience a fully asymptomatic course of infection vary with age. A. Analogous to Fig 3C in the main text: the probability that a single infected individual in any given age group triggers a local outbreak (grey bars) and the weighted average local outbreak probability $P$ (black horizontal line). B. Analogous to Fig 4D in the main text: partial reductions in 'school', 'work' and 'other' contacts, and the resulting reductions in the average local outbreak probability $P$ (solid red, dashed blue and dotted green lines respectively). C. Analogous to Fig 5D in the main text: the effect of reducing 'other' contacts on the average local outbreak probability when 'school' and 'work' contacts are not reduced at all (dotted line) and when 'school' and 'work' contacts are reduced by 100% (solid line). D. Analogous to Fig 6E in the main text: the age-dependent probability of a local outbreak with enhanced surveillance of both symptomatic and nonsymptomatic infected hosts ($\rho_k = 1/2$ days$^{-1}$ and $\sigma_k = 1/7$ days$^{-1}$), without contact-reducing NPIs (purple bars and solid line). Pale grey bars and black dash-dotted line represent the local outbreak probabilities without any contact-reducing NPIs or enhanced surveillance (as in Fig S9A).

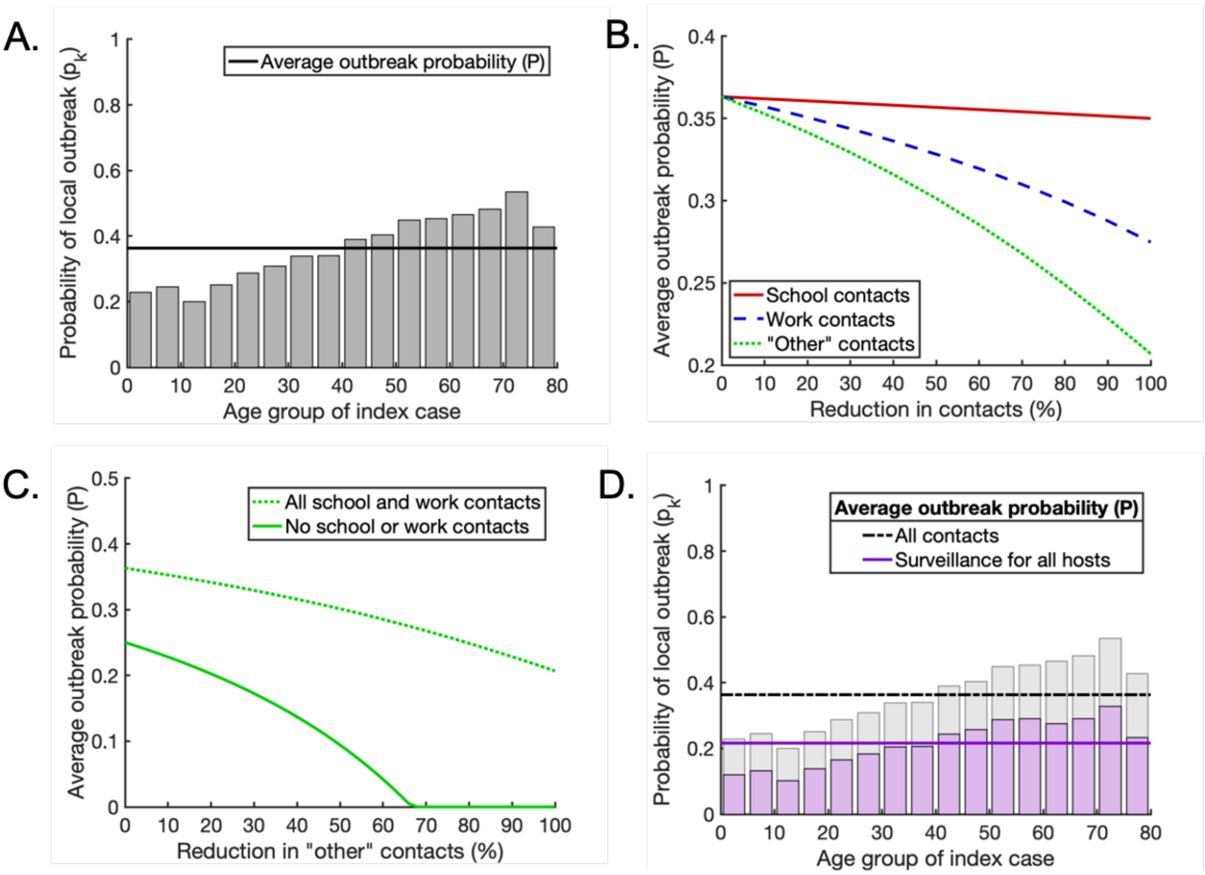


**Fig S10. Scenario C: The effect of increasing the proportion of infections that arise from presymptomatic hosts from $K_p = 0.489$ (baseline value) to $K_p = 0.75$.** The proportions of infections arising from symptomatic and asymptomatic hosts are adjusted so that they remain in the same ratio as in the baseline case. For scenario C, both susceptibility to infection and the proportion of hosts who experience a fully asymptomatic course of infection vary with age. A. Analogous to Fig 3C in the main text: the probability that a single infected individual in any given age group triggers a local outbreak (grey bars) and the weighted average local outbreak probability *P* (black horizontal line). B. Analogous to Fig 4D in the main text: partial reductions in 'school', 'work' and 'other' contacts, and the resulting reductions in the average local outbreak probability *P* (solid red, dashed blue and dotted green lines respectively). C. Analogous to Fig 5D in the main text: the effect of reducing 'other' contacts on the average local outbreak probability when 'school' and 'work' contacts are not reduced at all (dotted line) and when 'school' and 'work' contacts are reduced by 100% (solid line). D. Analogous to Fig 6E in the main text: the age-dependent probability of a local outbreak with enhanced surveillance of both symptomatic and nonsymptomatic infected hosts ($\rho_k = 1/2$ days$^{-1}$ and $\sigma_k = 1/7$ days$^{-1}$), without contact-reducing NPIs (purple bars and solid line). Pale grey bars and black dash-dotted line represent the local outbreak probabilities without any contact-reducing NPIs or enhanced surveillance (as in Fig S10A).

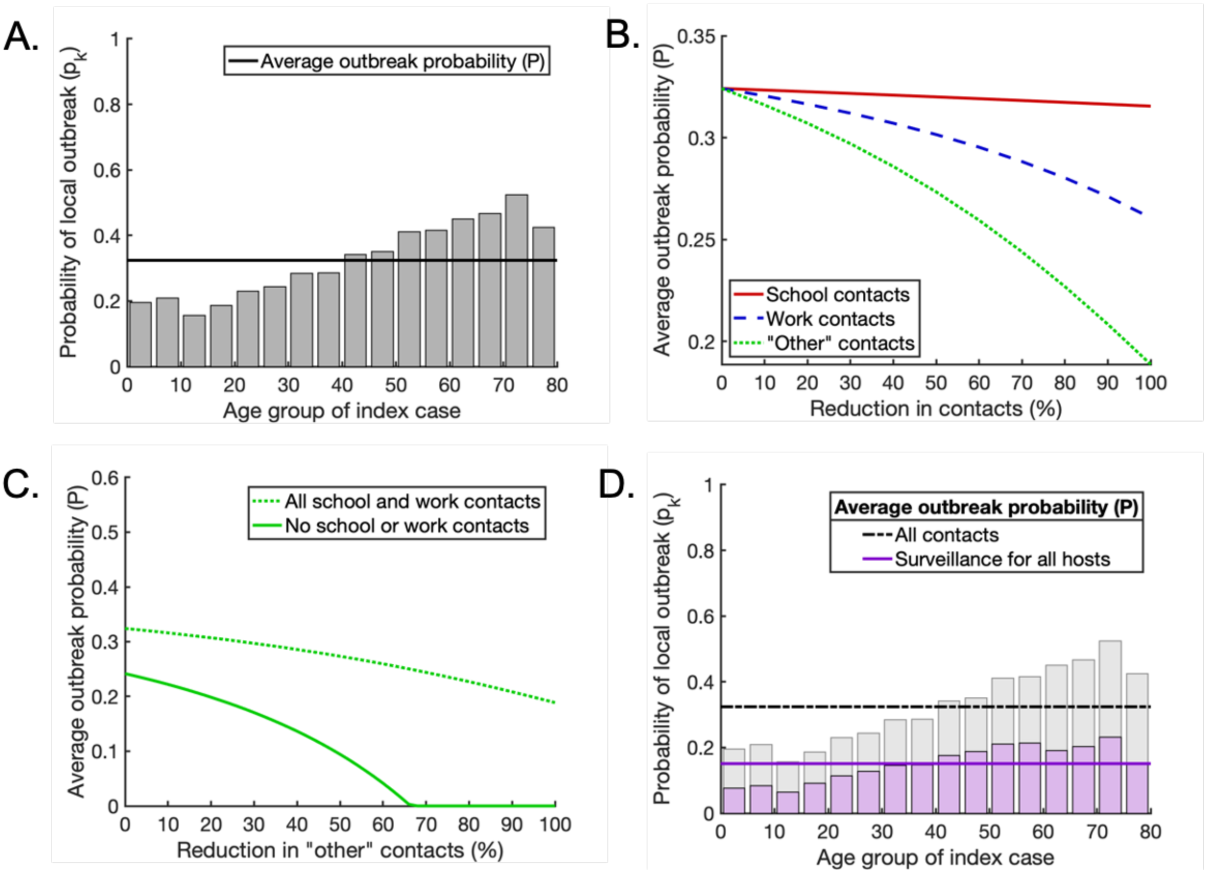


**Fig S11. Scenario C: The effect of reducing the proportion of infections that arise from asymptomatic hosts from $K_a = 0.106$ (baseline value) to $K_a = 0.01$.** The proportions of infections arising from symptomatic and presymptomatic hosts are adjusted so that they remain in the same ratio as in the baseline case. For scenario C, both susceptibility to infection and the proportion of hosts who experience a fully asymptomatic course of infection vary with age. A. Analogous to Fig 3C in the main text: the probability that a single infected individual in any given age group triggers a local outbreak (grey bars) and the weighted average local outbreak probability $P$ (black horizontal line). B. Analogous to Fig 4D in the main text: partial reductions in ‘school’, ‘work’ and ‘other’ contacts, and the resulting reductions in the average local outbreak probability $P$ (solid red, dashed blue and dotted green lines respectively). C. Analogous to Fig 5D in the main text: the effect of reducing ‘other’ contacts on the average local outbreak probability when ‘school’ and ‘work’ contacts are not reduced at all (dotted line) and when ‘school’ and ‘work’ contacts are reduced by 100% (solid line). D. Analogous to Fig 6E in the main text: the age-dependent probability of a local outbreak with enhanced surveillance of both symptomatic and nonsymptomatic infected hosts ($\rho_k = 1/2$ days$^{-1}$ and $\sigma_k = 1/7$ days$^{-1}$), without contact-reducing NPIs (purple bars and solid line). Pale grey bars and black dash-dotted line represent the local outbreak probabilities without any contact-reducing NPIs or enhanced surveillance (as in Fig S11A).

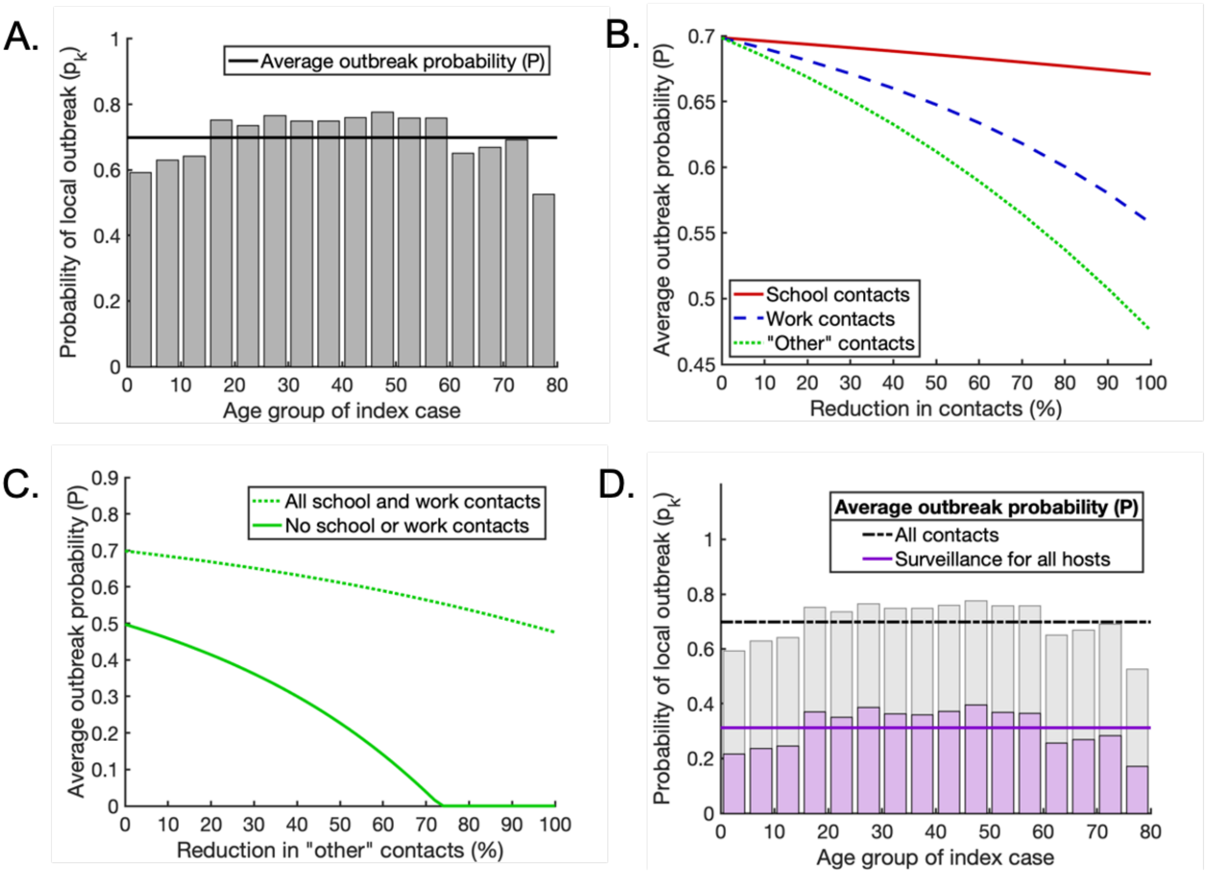


**Fig S12. Scenario C: The effect of increasing the proportion of infections that arise from asymptomatic hosts from $K_a = 0.106$ (baseline value) to $K_a = 0.5$.** The proportions of infections arising from symptomatic and presymptomatic hosts are adjusted so that they remain in the same ratio as in the baseline case. For scenario C, both susceptibility to infection and the proportion of hosts who experience a fully asymptomatic course of infection vary with age. A. Analogous to Fig 3C in the main text: the probability that a single infected individual in any given age group triggers a local outbreak (grey bars) and the weighted average local outbreak probability $P$ (black horizontal line). B. Analogous to Fig 4D in the main text: partial reductions in 'school', 'work' and 'other' contacts, and the resulting reductions in the average local outbreak probability $P$ (solid red, dashed blue and dotted green lines respectively). C. Analogous to Fig 5D in the main text: the effect of reducing 'other' contacts on the average local outbreak probability when 'school' and 'work' contacts are not reduced at all (dotted line) and when 'school' and 'work' contacts are reduced by 100% (solid line). D. Analogous to Fig 6E in the main text: the age-dependent probability of a local outbreak with enhanced surveillance of both symptomatic and nonsymptomatic infected hosts ($\rho_k = 1/2 \text{ days}^{-1}$ and $\sigma_k = 1/7 \text{ days}^{-1}$), without contact-reducing NPIs (purple bars and solid line). Pale grey bars and black dash-dotted line represent the local outbreak probabilities without any contact-reducing NPIs or enhanced surveillance (as in Fig S12A).

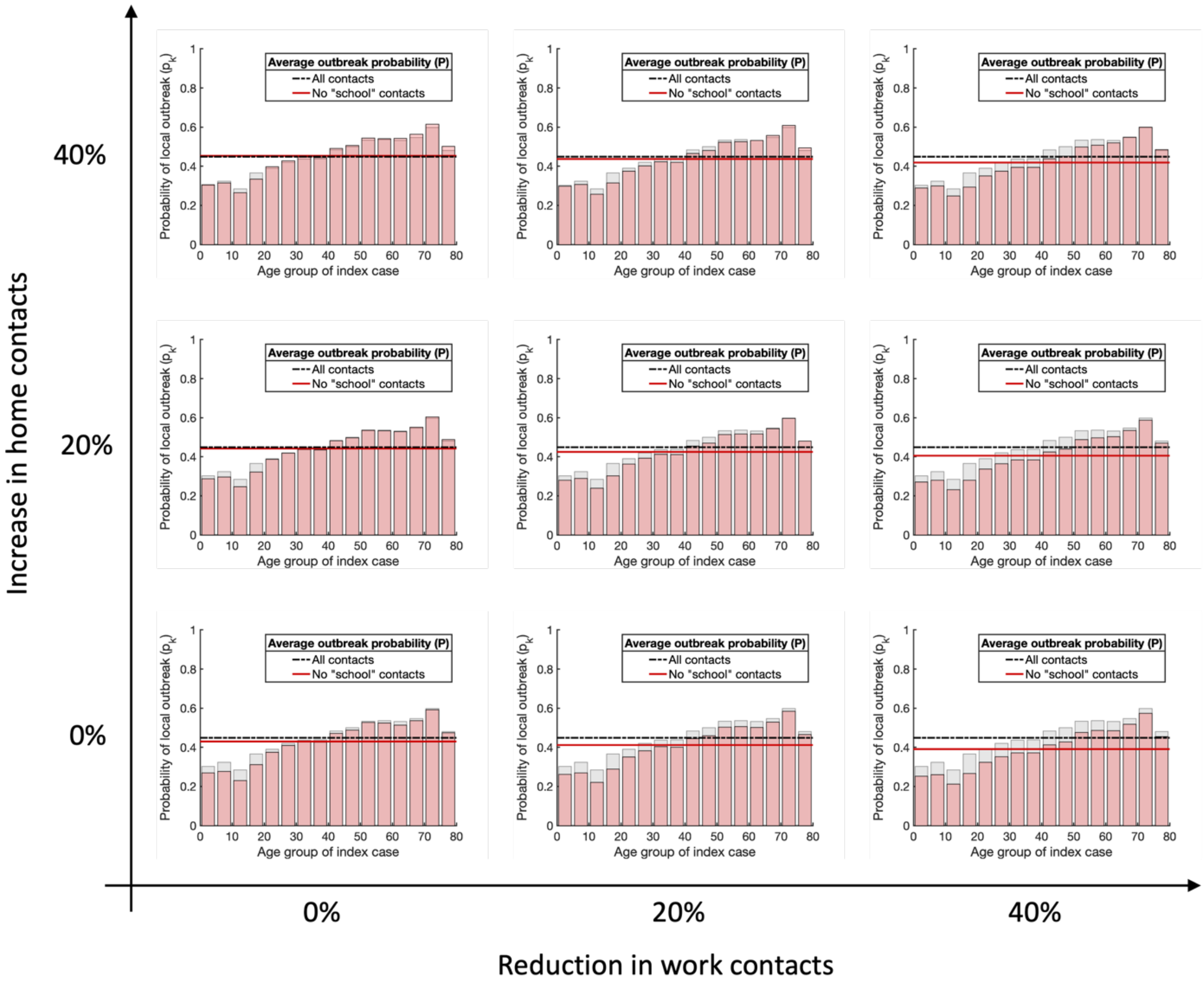


**Fig S13. Scenario C: The effects of school closures on the age-dependent local outbreak probability, allowing for possible secondary effects on 'work' and 'home' contacts.** In each panel the age-dependent local outbreak probability is shown in the absence of all 'school' contacts, allowing also for a specified reduction in 'work' contacts and increase in 'home' contacts that may occur as a result of school closures. Columns left to right represent a 0%, 20% and 40% reduction in 'work' contacts respectively. Rows bottom to top represent a 0%, 20% and 40% increase in 'home' contacts respectively. In every case, we assume that 'other' contacts are unaffected and remain as shown in Fig 2F of the main text. Although the shape of the age-dependent risk profile in the absence of 'school' contacts is robust to these changes in 'work' and 'home' contacts, the weighted average local outbreak probability $P$ (indicated by the solid red line in every case) varies. In particular, if the increase in 'home' contacts occurring as a result of school closures is high enough, this may counteract the benefits of reduced 'school' contacts. These results support our conclusion that school closures are unlikely to have a substantial impact on SARS-CoV-2 transmission when applied as the sole NPI.

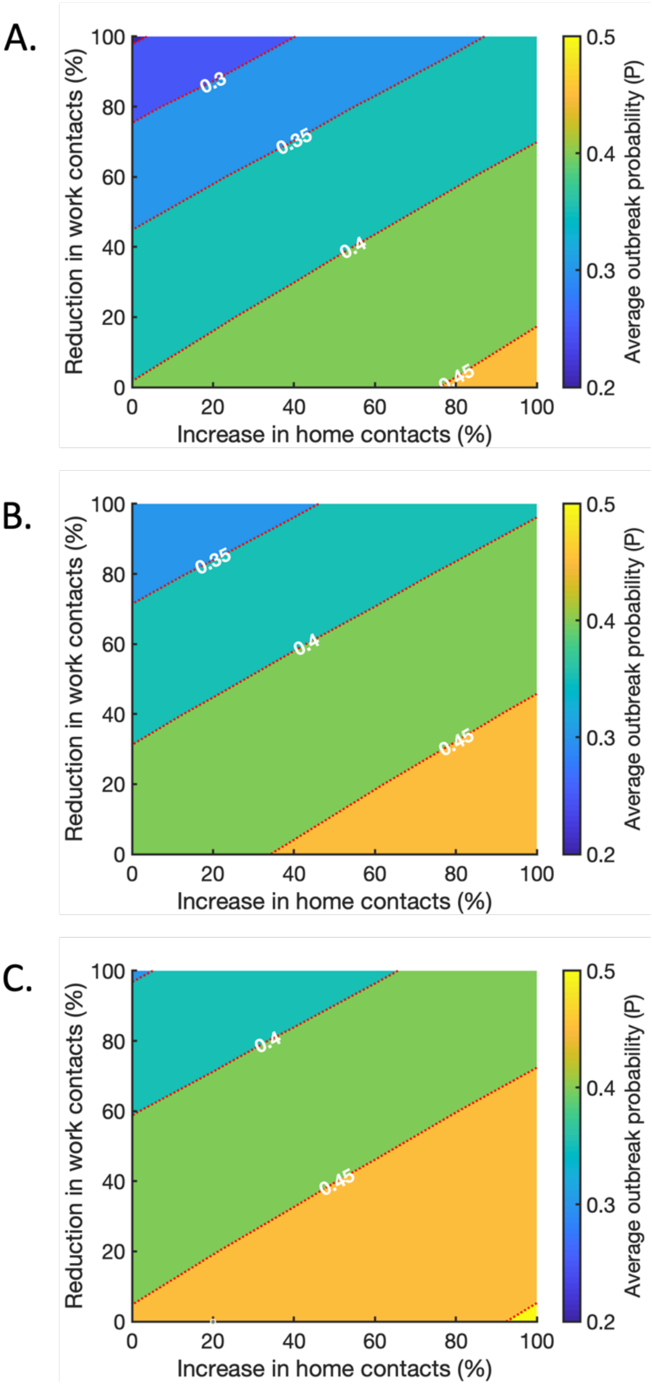


**Fig S14. Scenario C: The effects of school closures on the age-dependent local outbreak probability, allowing for possible secondary effects on 'work', 'home' and 'other' contacts.** A. The effect of increasing 'home' contacts and reducing 'work' contacts on the weighted average probability of a local outbreak (*P*), when 'other' contacts are reduced by 20% and 'school' contacts are removed entirely. Red dotted lines indicate contours along which the local outbreak probability is constant. B. The analogous figure to A, but with a 0% change in 'other' contacts compared to the baseline level. C. The analogous figure to A, but with a 20% increase in 'other' contacts.
